# Heterologous CoronaVac plus Ad5-nCOV versus homologous CoronaVac vaccination among elderly: a phase 4, non-inferiority, randomized study

**DOI:** 10.1101/2022.06.03.22275983

**Authors:** Pengfei Jin, Jingxin Li, Xiling Guo, Jinbo Gou, Lihua Hou, Zhizhou Song, Tao Zhu, Hongxing Pan, Jiahong Zhu, Fengjuan Shi, Pan Du, Haitao Huang, Jingxian Liu, Hui Zheng, Xue Wang, Yin Chen, Peng Wan, Shipo Wu, Xuewen Wang, Xiaoyu Xu, Wei Chen, Fengcai Zhu

## Abstract

**Importance:** People over 60 developed less protection after two doses of inactivated COVID-19 vaccine than younger people. Heterologous vaccination might provide greater immunity and protection against variants of concern.

**Objective:** To assess the safety and immunogenicity of a heterologous immunization with an adenovirus type 5-vectored vaccine (Convidecia) among elderly who were primed with an inactivated vaccine (CoronaVac) previously.

**Design:** An observer-blind, randomized (1:1) trial, conducted from August 26 to November 13, 2021.

**Setting:** A single center in Jiangsu Province, China.

**Participants:** 299 participants aged 60 years and older, of them 199 primed with two doses of CoronaVac in the past 3-6 months and 100 primed with one dose of CoronaVac in the past 1-2 months.

**Intervention:** Convidecia or CoronaVac as boosting dose

**Main Outcomes and Measures:** Geometric mean titers (GMTs) of neutralizing antibodies against wild-type SARS-CoV-2, and Delta and Omicron variants 14 days post boosting, and adverse reactions within 28 days.

**Results:** In the three-dose regimen cohort (n=199; mean (SD) age, 66.7 (4.2) years; 74 (37.2%) female), 99 and 100 received a third dose of Convidecia (group A) and CoronaVac (group B), respectively. In the two-dose regimen cohort (n=100; mean (SD) age, 70.5 (6.0) years; 49 (49%) female), 50 and 50 received a second dose of Convidecia (group C) and CoronaVac (group D), respectively. GMTs of neutralizing antibodies against wild-type SARS-CoV-2 at day 14 were 286.4 (95% CI: 244.6, 335.2) in group A and 48.2 (95% CI: 39.5, 58.7) in group B, with GMT ratio of 6.2 (95% CI: 4.7, 8.1), and 70.9 (95% CI: 49.5, 101.7) in group C and 9.3 (95% CI: 6.2, 13.9) in group D, with GMT ratio of 7.6 (95% CI: 4.1, 14.1). There was a 6.3-fold (GMTs, 45.9 vs 7.3) and 7.5-fold (32.9 vs 4.4) increase in neutralizing antibodies against Delta and Omicron variants in group A, respectively, compared with group B. However, there was no significant difference between group C and group D. Both heterologous and homologous booster immunizations were safe and well tolerated.

**Conclusions and Relevance:** Heterologous prime-boost regimens with CoronaVac and Convidecia induced strong neutralizing antibodies in elderly, which was superior to that induced by the homologous boost, without increasing safety concerns.

**Trial Registration:** ClinicalTrials.gov NCT04952727

**Key Points:** 

**Question:** Does a heterologous immunization with recombinant adenovirus type 5-vectored vaccine (Convidecia) produced a non-inferior or superior response of neutralizing antibodies among elderly primed with two doses of inactivated COVID-19 vaccine (CoronaVac), compared to the homologous boosting

**Findings:** In this randomized clinical trial, a heterologous third dose of Convidecia resulted in a 6.2-fold (geometric mean titers: 286.4 vs 48.2), 6.3-fold (45.9 vs 7.3) and 7.5-fold (32.9 vs 4.4) increase in neutralizing antibodies against wild-type strain, Delta and Omicron variants 14 days post boosting, respectively, compared to the homologous boost with CoronaVac

**Meaning:** Heterologous prime-boost regimens with CoronaVac and Convidecia induced strong neutralizing antibodies in elderly, which was superior to that induced by the homologous boosting.

## Introduction

Currently, the Omicron variant has become the dominant SARS-CoV-2 virus circulating globally, which is raising concerns because of its increased transmissibility and the potential to evade neutralizing antibodies elicited by COVID-19 vaccines^1-5^. In contrast to younger adults, persons 60 years of age or older face the highest rates of hospitalization and death associated with SARS-CoV-2 infection. Therefore, older adults are identified as a priority group for COVID-19 primary and boosting vaccinations, particularly for those primed with inactivated COVID-19 vaccines ^6,7^.

Inactivated COVID-19 vaccine CoronaVac (Sinovac) is one of the most widely used vaccines against SARS-CoV-2 worldwide, especially in low- and middle-income countries, which has been authorized to use by 55 countries^8^. Studies from Hong Kong and Brazil^9,10^ showed that primary vaccination with two doses of CoronaVac provide little protection against mild or moderate COVID-19 diseases associated with Omicron variant, and offered moderate protection against severe COVID-19 and death across all age groups. However, a third dose of BNT162b2 had higher vaccine effectiveness against hospitalization and death among individuals aged ≥75 years who had received two doses of CoronaVac, compared with the homologous boosting (79.9% vs 54.6%)^10^.

Convidecia (CanSino), is a recombinant adenovirus type-5 COVID-19 vaccine, which has been approved in 10 countries, including China, Argentina, Chile, Mexico, Pakistan, Indonesia, Malaysia and et al^8^. World Health Organization (WHO) issued an emergency use listing for Convidecia on 19 May 2022^11,12^. In a previous study, we found that heterologous boosting of CoronaVac with Convidecia produced higher neutralizing antibodies compared to the homologous boosting with CoronaVac did in adults aged 18-59 years, with GMT ratios of heterologous group vs. homologous group around 4.3∼5.9^13^. These data have supported that a heterologous boosting immunization strategy been proposed for use in adults by health agencies in China. However, the study of a heterologous boosting with Convidecia in the older population whom were primed with CoronaVac has not been revealed.

Here, we report the safety and immunogenicity of a heterologous immunization with Convidecia in the individuals aged over 60 years who were primed with CoronaVac before.

## Methods

### Trial design and participants

We conducted an observer-blind, randomized (1:1), parallel-controlled trial to assess the safety and immunogenicity of heterologous immunization with Convidecia vs. homologous immunization with CoronaVac in elderly. Participants were recruited in Lianshui County, Jiangsu Province, China. The trial protocol and informed consent form were reviewed and approved by the ethics committee of the Jiangsu Provincial Center for Disease Control and Prevention. This trial was conducted following the principles of the Declaration of Helsinki and Good Clinical Practice Guidelines. The complete trial protocol is available in Supplement 1.

Participants aged 60 years or older, who had completed two doses of CoronaVac in the past 3-6 months or one dose of CoronaVac in the past 1-2 months, were recruited. Participants who had a confirmed history of COVID-19 or SARS-CoV-2 infection, severe or unstable comorbidities, allergic reactions or hypersensitivity to vaccine components were excluded. A full list of the inclusion and exclusion criteria is provided in the protocol.

### Randomization and masking

We used an interactive web-based response-randomization system for stratified randomization. Eligible participants who had received two doses of CoronaVac were randomized (1:1) to receive a third dose of Convidecia (group A, heterologous boosting) or CoronaVac (group B, homologous boosting), while participants who had been primed with one dose of CoronaVac were randomized equally to receive a second dose of Convidecia (group C, heterologous dose) or CoronaVac (group D, homologous dose). Randomization lists were generated by an independent statistician using SAS (version 9.4), with a block size of 10.

The unblinded staffs who prepared and administered vaccination were aware of the treatment allocation, but were not allowed to involve in any other trial procedures or to reveal this information to any participants or other investigators. Blinding was maintained by preparing vaccines out of sight of participants and other investigators and concealing the syringes with a label of randomization number.

### Procedures

For both CoronaVac and Convidecia, the administration is via 0.5mL intramuscular injection into the upper arm. Participants were asked to stay at least 30 min after vaccination for any immediate adverse reactions and were instructed to record any solicited or unsolicited adverse events up to day 28 after vaccination on paper diary cards. Serious adverse events (SAEs) reported by participants were documented throughout the 6-month trial duration.

Blood samples were collected for immunogenicity assessments on day 0 before the vaccination and on day 14, and 28 after the vaccination. The specific methods have been previously described^13^. Neutralizing antibodies against live SARS-CoV-2 virus were measured by using a cytopathic effect-based microneutralization assay with a wild-type SARS-CoV-2 virus isolate BetaCoV/Jiangsu/JS02/2020 (GISAID EPI_ISL_411952), and a SARS-CoV-2 Delta (B.1.617.2) variant isolate hCoV-19/China/JS07/2021(GISAID EPI_ISL_4515846), and Omicron (BA.1.1) variant isolate hCoV-19/Jiangsu/JS01/2022(GISAID EPI_ISL_12511653) in Vero-E6 cells. Serum dilution for the microneutralization assay started from 1:4 to 1:512. Serum dilutions were mixed with the same volume of viral solution to achieve a final concentration of 100 TCID50 per well. The reported titer was the reciprocal of the highest sample dilution that protected at least 50% of cells from cytopathic effects. IgG binding antibody concentrations against receptor-binding domain (RBD) was detected by an indirect enzyme-linked immunosorbent assay (ELISA) (Vazyme Biotech Co., Ltd). The WHO international standard for anti-SARS-CoV-2 immunoglobulin (NIBSC code 20/136) was used side by side as a reference with the serum samples measured in this study for calibration and harmonization of both the microneutralization and ELISA assays.

Peripheral blood monouclear cells (PBMCs) from blood samples of the first 50 and 30 participants in the three-dose and two-dose regimen cohorts, respectively, were collected to evaluate cellular immunity. Th1-secreted cytokines (IFN-γ and TNF-α) and Th2-secreted cytokines (IL-4, IL-5, and IL-13) were detected by ELISpot assay (Mabtech, Stockholm, Sweden) after the PBMCs been stimulated by using the overlapping peptide pool of spike glycoprotein.

### Outcomes

The primary endpoints were the GMT of neutralizing antibodies to wild-type SARS-CoV-2 at day 14 after vaccination. Immunological secondary outcomes included neutralizing antibodies to wild-type SARS-CoV-2 at day 28, and anti-receptor binding domain (RBD) binding IgG antibody and neutralization titers against SARS-CoV-2 virus variants of concern at days 14 and 28. Spike-specific T-cell responses were measured by ELISpot before and 14 days after vaccination. Safety outcomes included adverse reactions within 28 days after vaccination and SAEs within 6 months

We also explored the effects of age in the populations receiving heterologous boost with Convidecia or homologous boost with CoronaVac as a post-hoc evaluation, by comparing the neutralizing antibody responses in the older participants with those observed in the young adults aged 18-59 years in a previous study (NCT04892459)^13^.

### Sample size

The sample size was calculated based on the assumption that a heterologous boost would elicit at least a non-inferior or superior response of neutralizing antibodies to the homologous boost (Group A vs Group B) did and was performed by using Power Analysis and Sample Size software (version 11.0.7). We assumed that the GMT of neutralizing antibodies was approximately 1:40 at the enrollment (3-6 months after two doses of CoronaVac), while GMTs on day 14 post boost were expected to reach 1:80 for participants receiving a homologous boost dose of CoronaVac and 1:160 for those receiving a heterologous boost dose of Convidecia, respectively. The standard deviation of the GMTs for the two groups was estimated to be 4. A total of 100 participants per group will provide more than 99% power to identify the non-inferiority of log-transformed neutralization titers at a non-inferiority margin of 0.67 and at least 90% power to detect the superiority of the heterologous boost. The probability of observing at least one specific adverse event with an incidence of 2% in each group of 100 participants was 86.7%. The heterologous vaccination after one dose of CoronaVac was explored (groups C and D), but was not considered as the primary target immunization schedule. Therefore, half of the sample size for groups A and B (50 participants per group) was applied for groups C and D without a power calculation.

### Statistical analysis

The primary immunogenicity analysis was done in the per-protocol cohort, including all participants who were injected and for whom serological samples were available on day 14 post boost vaccination. Antibody levels against SARS-CoV-2 were log-transferred and then calculated for GMTs, geometric mean fold increases (GMFIs). Antibody titers measured below the detection limit were replaced by a value equal to half of the lowest limit. The proportion of participants with at least a 4-fold increase was presented with 95% CIs. The GMT ratio was calculated as the antilogarithm of the difference between the mean of the log-transformed titer of the heterologous group vs the corresponding homologous group.

Participants who received a study vaccine were included in the safety analysis. The safety data were presented as numbers and percentages of participants who have suffered at least one local or systemic adverse reaction within 28 days after the vaccination. We analyzed categorical data with the χ2 test or Fisher’s exact test, log-transformed antibody titers with the t-test, and data that did not follow a normal distribution with the Wilcoxon rank-sum test. Correlations between neutralizing antibodies against SARS-CoV-2 and anti-RBD binding IgG antibodies were evaluated by Pearson correlation coefficients. To explore the association between age as a categorical variable and log-transformed neutralizing antibody titers, a multivariate linear regression was fitted, adjusting for vaccine group, vaccination interval, baseline antibody titers and gender. Statistical analyses were performed using R (version 4.0.5).

## Results

### Study participants

From August 26 to November 13, 2021, 326 volunteers were recruited and screened for eligibility and 299 eligible participants were enrolled. Of them, 199 participants primed with two doses of CoronaVac were included in a three-dose regimen cohort, with 99 in group A and 100 in group B. 100 participants primed with one dose of CoronaVac were included in a two-dose regimen cohort, with 50 in group C and 50 in group D, respectively (Figure 1). All the participants completed a 28-day follow-up to assess safety. We obtained blood samples from 299 participants on day 0 before the vaccination, and from 293 participants on day 14, and from 288 participants on day 28, respectively.

**Figure 1.**
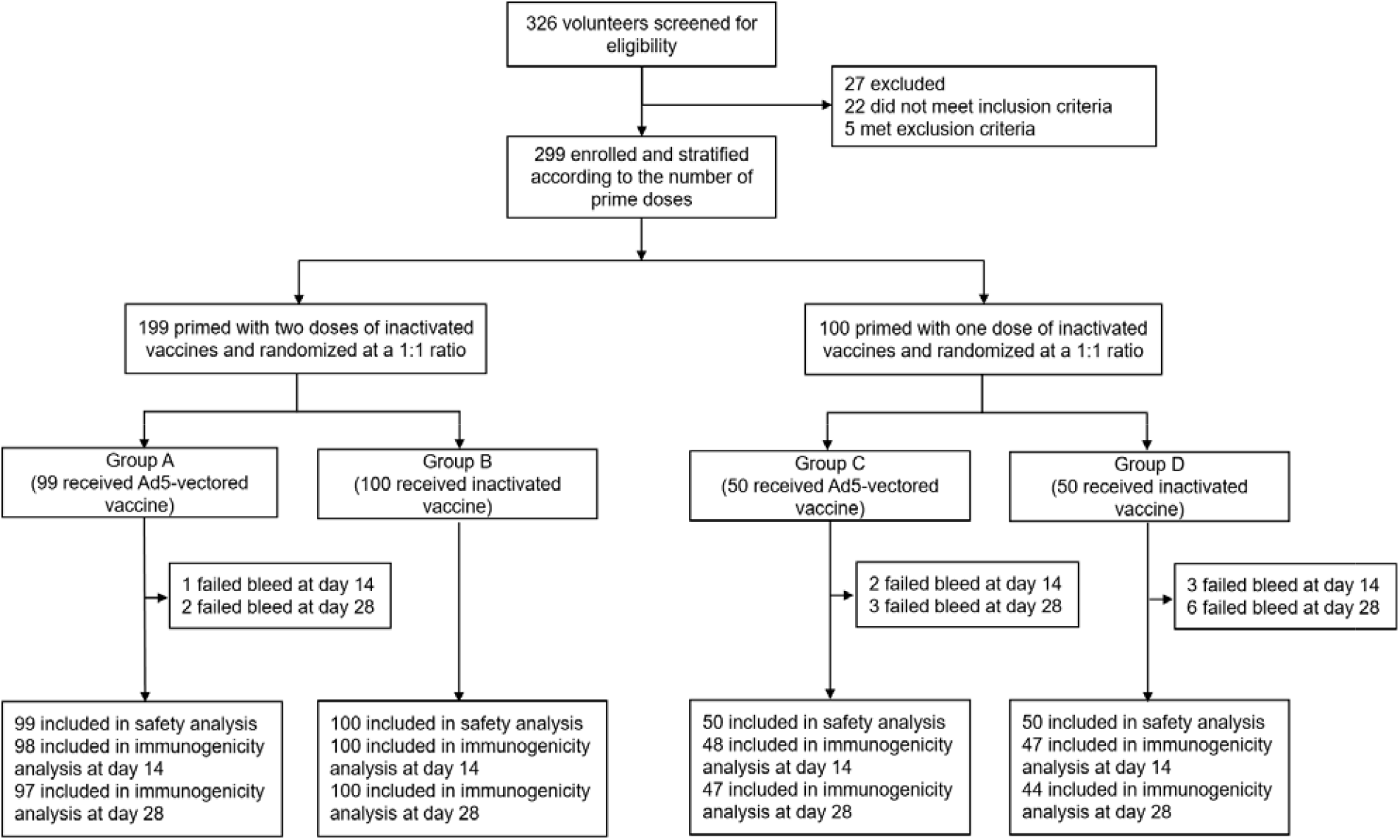
CONSORT Flowchart. Six participants failed to bleed at 14 days, with one participant in group A, two in group C and three in group D. Eleven participants failed to bleed at 28 days, with two participant in group A, three in group C and six in group D. All participants completed 6 months of safety follow-up

In the three-dose regimen cohort, the average age of participants was 66.7 years, and 74 of 199 (37.2%) were female, with a median interval of 4.9 (interquartile range (IQR): 4.7, 5.0) since the second dose. In the two-dose regimen cohort, the average age of participants was 70.5 years, and 49 of 100 participants (49%) were female, with a median interval of 1.1 (IQR: 1.1, 1.1) months since receiving the first dose. The demographic characteristics of the participants were comparable between the heterologous and homologous groups (Table 1).

**Table 1.** Baseline characteristics of the participants.

|  | **Group A**<br>(Corona Vac/Corona Vac+<br>Convidecia, n=99) | **Group B**<br>(Corona Vac/Corona Va+<br>Corona Vac, n=100) | **Group C**<br>(Corona Vac+<br>Convidecia, n=50) | **Group D**<br>(Corona Vac+<br>Corona Vac, n=50) |
| --- | --- | --- | --- | --- |
| **Age, years** |  |  |  |  |
| Mean<br>(SD) | 66.6 (4.6) | 66.8 (3.8) | 71.0 (6.2) | 69.9 (5.9) |
| Median<br>(IQR) | 66.0 (64.0,70.0) | 66.0 (64.0,69.0) | 72.0 (67.0,74.0) | 70.5 (66.0,73.0) |
| **Sex (%)** |  |  |  |  |
| Male | 65 (65.7%) | 60 (60.0%) | 28 (56.0%) | 23 (46.0%) |
| Female | 34 (34.3%) | 40 (40.0%) | 22 (44.0%) | 27 (54.0%) |
| **Time since the last priming dose of inactivated vaccine (months)** |  |  |  |  |
| Median<br>(IQR) | 4.9 (4.8, 5.0) | 4.9 (4.8, 5.1) | 1.2 (1.1, 1.2) | 1.1 (1.1, 1.2) |

Data are n (%) or mean ± SD. or median (IQR). n = number of participants. % = proportion of participants. SD=standard deviation. IQR=interquartile range.

### Neutralizing antibody responses against the SARS-CoV-2 virus

No detectable neutralizing antibodies against wild-type SARS-CoV-2 in serum were observed among the participants at the enrollment, so half of the lowest limit titer of 1:4 was assumed for baseline (Figure 2). Among participants whom were primed with two doses of CoronaVac, GMTs of neutralizing antibodies against wild-type SARS-CoV-2 increased to 286.4 (95% CI: 244.6, 335.2) in group A and 48.2 (95% CI: 39.5, 58.7) in group B 14 days post boost vaccination, with GMFIs of 143.2 (95% CI: 122.3, 167.6) and 24.1 (95% CI: 19.8, 29.4) from the baseline, respectively. Among participants whom were primed with one dose of CoronaVac, a heterologous second dose induced a GMT of neutralizing antibodies against wild-type SARS-CoV-2 of 70.9 (95% CI: 49.5, 101.7) in group C, while a homologous dose induced 9.3 (95% CI: 6.2, 13.9) in group D at day 14, with GMFIs of 35.5 (95% CI: 24.8, 50.8) and 4.6 (95% CI: 3.1, 6.9), respectively (Figure 2; eTable 1 in the Supplement 2).The GMT ratios of the heterologous vs. homologous groups were 6.2 (95% CI: 4.7, 8.1) for three-dose regimens and 7.6 (95% CI: 4.1, 14.1) for two-dose regimens (eTable 1 in the Supplement 2). At 28 days after the vaccination, the neutralizing antibodies against wild-type SARS-CoV-2 decreased slightly in all groups (Figure 2), but the level of which in heterologous group remained significantly higher than that in the homologous group (eTable 1 in the Supplement 2).

**Figure 2.**
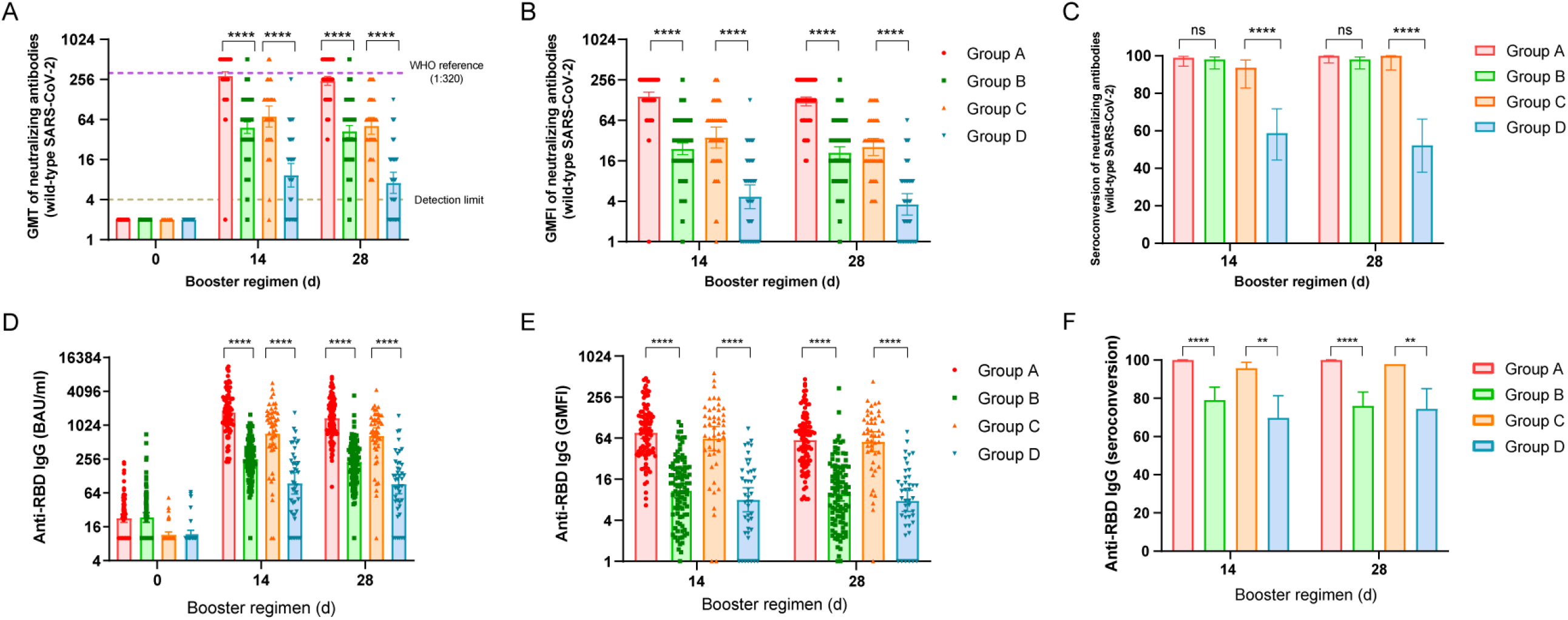
Neutralizing antibody titers against wild-type SARS-CoV-2 and RBD-specific IgG antibodies before and after boosting. GMTs of neutralizing antibodies against wild-type SARS-CoV-2 (A) or anti-RBD IgG antibodies (D); GMFI of neutralizing antibodies against wild-type SARS-CoV-2 (B) or anti-RBD IgG antibodies (E); Seroconversion of neutralizing antibodies against wild-type SARS-CoV-2 (C) or anti-RBD IgG antibodies (F). Error bars indicate 95% CIs. Seroconversion was defined as the proportion of participants with at least a fourfold increase of post-vaccination antibody levels compared to levels before the booster vaccination. Group A, primed with two doses of CoronaVac and given one dose of Convidecia (n=□98); Group B, primed with two doses of CoronaVac and given one dose of CoronaVac (n=□100); Group C, primed with one dose of CoronaVac and given one dose of Convidecia (n=□48); Group D, primed with one dose of CoronaVac and given one dose of CoronaVac (n=□47). n, the number of participants included in the per-protocol cohort. The WHO reference (1,000□IU/□ml) is equivalent to a live viral neutralizing antibody titer of 1:320 against wild-type SARS-CoV-2. *P* values result from a comparison between the two treatment groups using t-tests for log-transformed antibody titers or two-sided χ^2^ tests for categorical data (Group A versus Group B, and Group C versus Group D). For (C, F), the statistics are the proportions of participants with seroconversion after the vaccination. **P<□0.01, ****P<□0.0001, ns, representing P>0.05.

### RBD-specific antibodies levels

In line with the neutralizing antibodies against wild-type SARS-CoV-2, both heterologous and homologous boost immunizations induced significant increases in RBD-binding IgG levels at day 14 (Figure 2). However, the heterologous boosting elicited higher RBD-binding IgG GMCs than homologous boosting in both regimen cohorts, with 1731.9 (95% CI: 1451.6, 2066.4) in group A versus 253.8 (95% CI: 214.7, 300.1) in group B, and 724.0 (95% CI: 477.0, 1098.8) in group C versus 94.7 (95% CI: 60.7, 147.6) in group D, with P values <0.0001 (eTable 2 in the Supplement 2). Similarly, only a slight decrease in anti-RBD IgG antibody levels were observed at 28 days after vaccination (Figure 2).

Anti-RBD IgG antibody responses were predominantly IgG1 after the boost across the treatment groups, but heterologous dose induced higher IgG1 levels at day 14 than that homologous vaccine did in both regimen cohorts. Increased IgG3 and IgG2 levels were mild across four groups, but still the heterologous boost groups (groups A and C) induced higher antibody titers than the homologous boost groups (groups B and D) did. No increase of the levels of IgG4 after the vaccination was observed (eFigure 1 in the Supplement 2). At day 14, the mean IgG1/IgG4 ratios were 234.4 (95%CI: 193.1, 284.4) in group A, 39.3 (95% CI: 32.6, 47.4) in group B, 137.6 (95% CI: 97.6, 193.7) in group C, and 19.4 (95% CI: 12.3, 30.5) in group D, respectively.

### Neutralizing antibody responses against Delta and Omicron variants

None of the participants had detectable neutralizing antibodies against Delta or Omicron variants at the baseline. At day 14 after the vaccination, 50 (100.0%) participants in group A were seropositive of neutralizing titers against Delta variant, following by 40 (80.0%) in group B, 14 (58.3%) in group C and 7 (30.4%) in group D The GMT of neutralizing antibodies against Delta variant in group A was 45.9 (95% CI: 34.5, 61.0), which was higher than that in group B (7.3, 95% CI: 5.6, 9.5) (P<0.0001). While the neutralizing antibodies against Delta variant were not significantly different between group C and D (eTable 1 in the Supplement 2). However, compared with neutralizing titers against wild-type SARS-CoV-2, the neutralizing capability to Delta variant were reduced by 6.2-fold for in group A, 6.6-fold in group B, 14.1-fold in group C, and 2.3-fold in group D (Figure 3).

**Figure 3.**
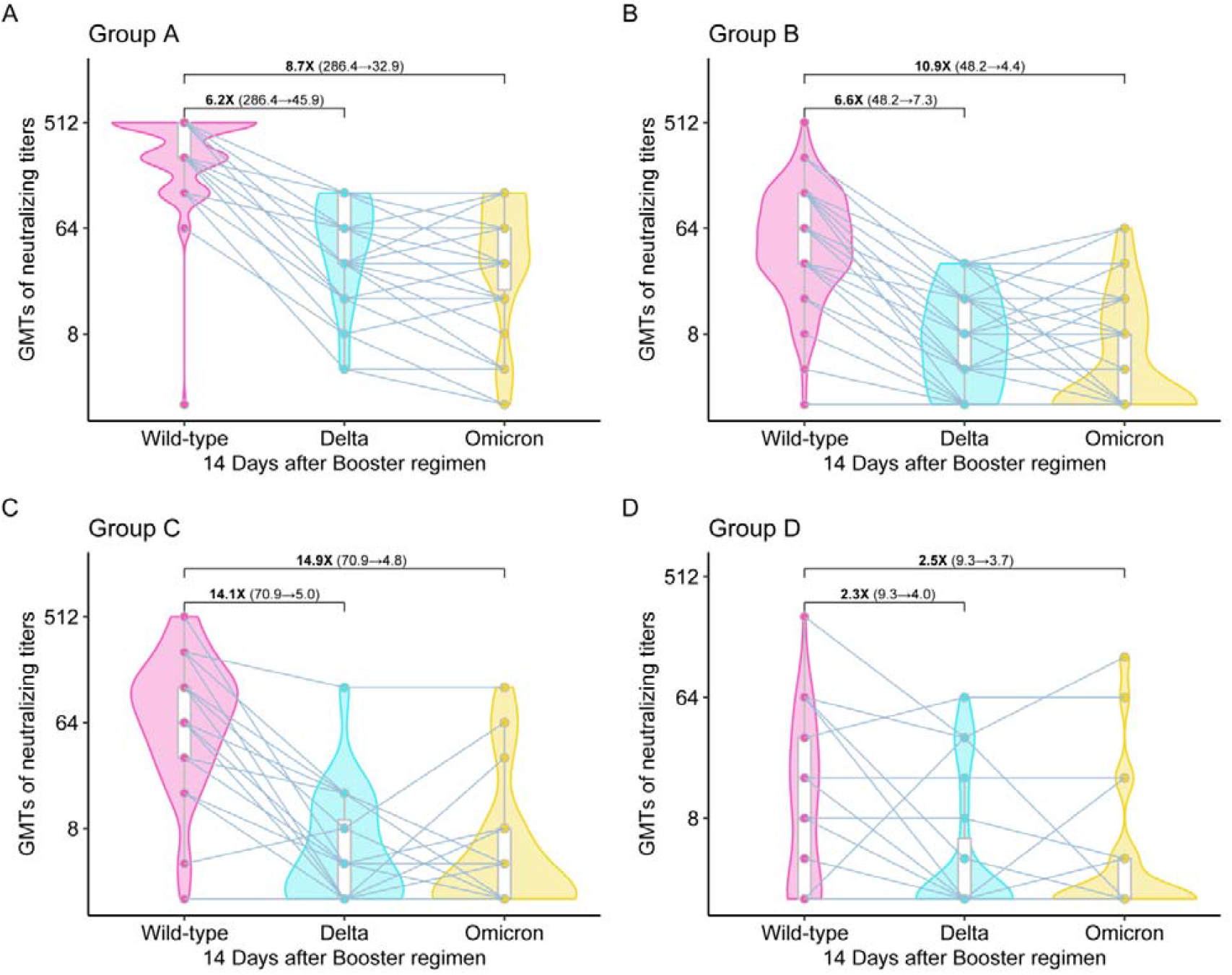
Comparison of neutralizing antibodies against SARS-CoV-2 wild-type, Delta variant, and Omicron variant 14 days after boosting. Box-violin plots of neutralizing antibody titers against Delta and Omicron variants compared to wild-type SARS-CoV-2 strain. Bold numbers indicate the geometric mean ratios neutralizing antibodies against wild-type relative to Delta or Omicron variants. Numbers in brackets represent geometric mean titers of neutralizing antibodies against wild-type and against Delta or Omicron variants. Group A, primed with two doses of CoronaVac and given one dose of Convidecia (n=□50); Group B, primed with two doses of CoronaVac and given one dose of CoronaVac (n=50); Group C, primed with one dose of CoronaVac and given one dose of Convidecia (n=□30); Group D, primed with one dose of CoronaVac and given one dose of CoronaVac (n=□30). All the paired data of neutralizing antibodies against wild-type SARS-CoV-2 and against Delta or Omicron variants from participants are included in the analysis. The discrepancies between the numbers of data points presented in the figures and the numbers of participants in the groups are due to the overlapping of the dots.

For the Omicron variant, seropositive rate at day 14 was 94.0% (47/50) in group A, 42.0% (21/50) in group B, 45.8% (11/24) in group C, and 30.4% (7/23) in group D, respectively. The neutralizing antibody GMT was also higher in group A (32.9, 95% CI: 23.1, 46.9) than in group B (4.4, 95% CI: 3.2, 5.9) (P<0.0001). However, no significant difference was shown between group C and group D for Omicron variant as well (eTable 1 in the Supplement 2). GMTs against Omicron variant was 8.7 times lower in group A and 10.9 times in group B, 14.9 times in group C, 2.5 times in group D, compared to that against wild-type SARS-CoV-2 (Figure. 3).

Strong correlations between anti-RBD IgG and neutralizing antibodies against wild-type SARS-CoV-2 were observed, with correlation coefficients of 0.711-0.827, among the four treatment groups. Correlations between anti-RBD IgG and neutralizing antibodies against Delta or Omicron variants remained strong for group A, with correlation coefficients of 0.812 and 0.707, but became weak or moderate for other groups (correlation coefficients of 0.269-0.692) (eFigure 2 in the Supplement 2). We found that age was negatively correlated to neutralizing antibodies against all three SARS-CoV-2 strains in two-dose regimens (groups C and D), but not in three-dose regimens (groups A and B) (eFigure 3 in the Supplement 2).

### Vaccine-induced T cell responses

We observed an increase in the levels of IFN-γ across all treatment groups 14 days post boosting (Figure 4A), with a median IFN-γ spot counts of 50 (IQR: 13, 118) in group A, 30 (IQR: 10, 85) in group B, 55 (IQR: 28, 203) in group C, and 20 (IQR: 5, 30) in group D per 10^6^ PBMCs, respectively. There were comparably high levels of TNF-α at baseline between treatment groups, and only increased slightly in groups B and D after boosting but not in groups A and C (Figure 4B). A slight increase was also observed in the levels of IL-4, IL-5 and IL-13 14 days post boosting, while there was not significant difference between treatment groups (Figure 4C-E). Nevertheless, the cytokine profile of Th1/Th2 suggested a Th1 skewing in both heterologous and homologous groups (Figure 4F).

**Figure 4.**
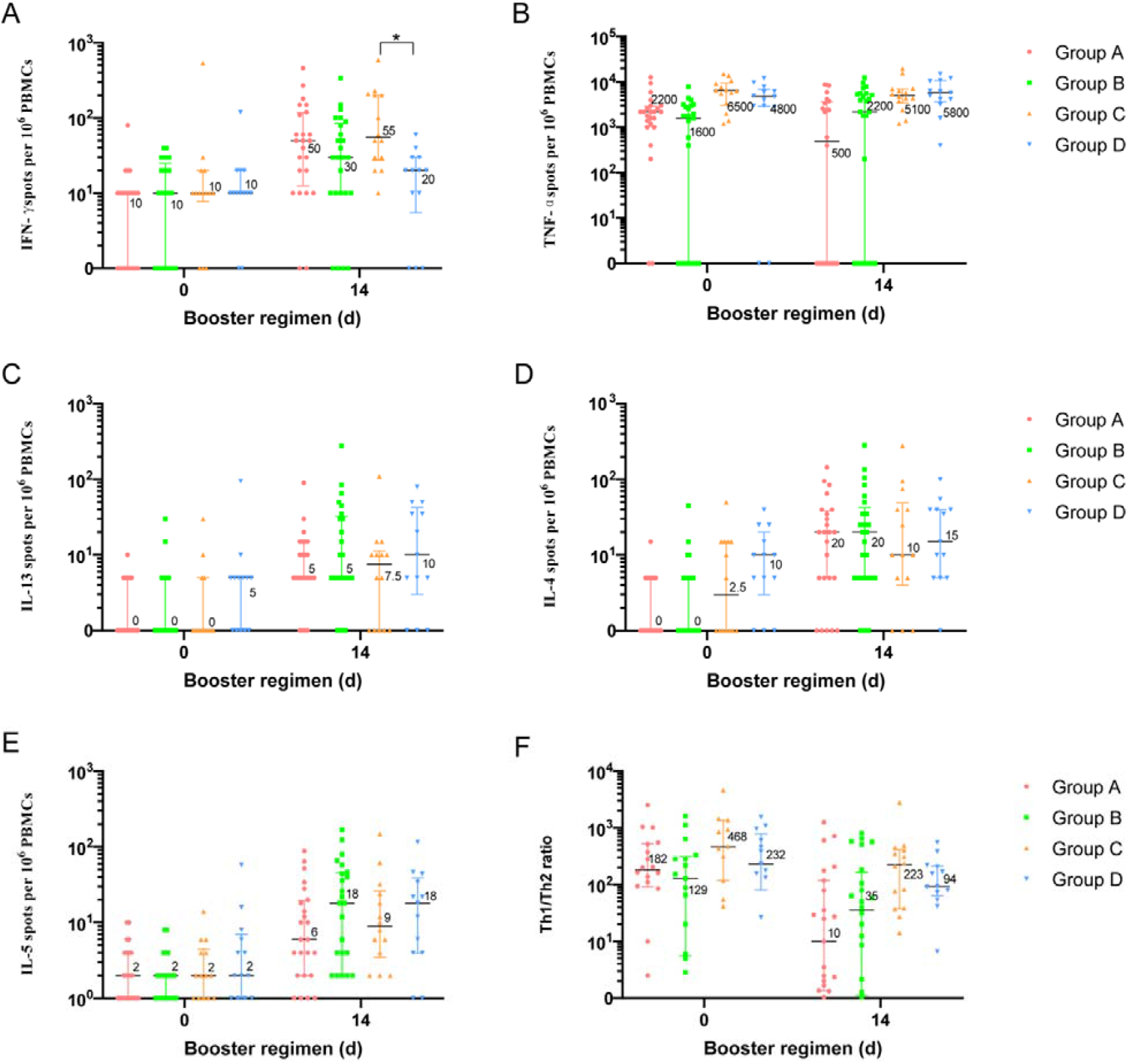
SARS-CoV-2 spike-specific T cell cytokine responses before and after boosting. The Th1/Th2 ratio (F) was calculated by summing IFN-γ (A) and TNF-α (B) cytokine levels and then dividing by the sum of IL-13 (C), and IL-4 (D), and IL-5 (E) cytokine levels. Data are the median of positive spot counts per 10^6^ PBMCs. Horizontal bars show the median, and error bars indicate interquartile range (IQR). Group A, primed with two doses of CoronaVac and given one dose of Convidecia (n=□25); Group B, primed with two doses of CoronaVac and given one dose of CoronaVac (n=□25); Group C, primed with one dose of CoronaVac and given one dose of Convidecia (n=□15); Group D, primed with one dose of CoronaVac and given one dose of CoronaVac (n=□15). Cytokine-secreting T cells were background corrected for unstimulated cells, and values lower than 0 were considered negative. *P* values result from a comparison between the two treatment groups using Wilcoxon rank-sum tests (group A versus group B, and group C versus group D). *P<□0.05.

### Safety

Either heterologous or homologous booster vaccination was safe and well tolerated in old participants aged equal to or over 60 years. Participants primed with two doses of CoronaVac reported similar frequent adverse reactions after receiving a heterologous dose of Convidecia or a homologous dose of CoronaVac. Adverse reactions within 28 days post boosting were reported in 8 (8.1%) of 99 recipients in group A and 4 (4.0%) of 100 recipients in group B. In participants primed with one dose of CoronaVac, adverse reactions were more frequent post boost vaccination with Convidecia compared with homologous boost (p=0.031), with the incidence rates of 16.0% (8/50) in group C and 2.0% (1/50) in group D, respectively. All adverse reactions were generally mild or moderate in severity, except for one severe pain at the injection site in group C (eTable 3 in the Supplement 2).

There were four SAEs occurred during the study, none of which are considered related to study vaccines (eTable 4 in the Supplement 2). There was no COVID-19 outbreak in trial site, and no COVID-19 cases were reported.

### Post-hoc analysis of the neutralizing antibody between the young and the old

Compared with the previous study with heterologous Convidecia boost after the CoronaVac primary series in adults aged 18-59 years, the neutralizing antibodies at the enrollment were even lower among elderly in this study. However, neutralizing antibodies against wild-type SARS-CoV-2 in old adults were higher than that in young adults at day 14 post boosting in the three-dose regimens (eFigure 4 in the Supplement 2). For the two-dose regimens, similar antibody titers against wild-type SARS-CoV-2 were seen between the young and the old. Multivariate linear regression showed that age had no significant effect on neutralizing antibodies (against SARS-CoV-2 wild-type, Delta or Omicron variants) 14 days after boosting for three-dose regimens, but was negatively correlated with antibody titers against Delta or Omicron variants in two-dose regimens (eTable 5 in the Supplement 2).

## Discussion

Our findings show that heterologous vaccination with Convidecia following primary immunization series of CoronaVac provided a substantial increase in antibody responses against wild-type SARS-CoV-2 in adults aged 60 years and older, which are superior to homologous vaccination of CoronaVac did, without increasing safety concerns. Participants who received the heterologous Convidecia in the third-dose regimen cohort had the highest levels of neutralizing antibodies, followed by those received the heterologous Convidecia in the two-dose regimen cohort and those received three or two-dose of CoronaVac. Of note, a heterologous third dose of Convidecia also markedly increased neutralizing antibodies to the Delta and Omicron variants in older adults who have been primed with two doses of CoronaVac, respectively.

The magnitude of the immune boost was greater with Convidecia compared with the homologous regimen, which was similar to recent findings following boosting with Ad26.COV2-S or ChAdOx1 nCoV-19 after two priming doses of CoronaVac^14^. A real-world study of 11.2 million persons aged 16 years and older in Chile showed also that heterologous booster dose (BNT162b2 or ChAdOx1 nCoV-19), in individuals with a complete primary vaccination schedule with CoronaVac, provided higher vaccine effectiveness against symptomatic COVID-19 than a homologous booster with 93.2% for a ChAdOx1 nCoV-19 booster and 96.5% for a BNT162b2 booster vs. 78.8% for CoronaVac, during the predominant circulating of Delta variant ^15^.

People older than 60 years have an increasing risk of severe illness and death from COVID-19, especially for those with underlying chronic conditions. The immune responses to vaccines are usually reduced in older adults due to immune senescence. In this study, we observed comparable or even slightly higher levels of the neutralizing antibodies against wild-type SARS-CoV-2 in older adults receiving three-dose regimens compared with those in adults aged 18-59 years previously reported^13^. In our study, this observation is probably biased by a longer interval between booster dose and primary vaccination in the older adults study than that in the younger adults study (the media of the intervals, 4.9 vs 3.3 months). Because the maturity of memory B cells take time, and the longer interval between the prime and the boost generally results more robust antibody responses^16,17^. In addition, the effect of age on neutralizing antibodies in elderly population could be underestimated, due to a too-small number of adults 80 years or older in our study (2.3% (7/299) of the study population).

Low neutralizing antibody responses against the Omicron variant have been observed in individuals receiving two doses of CoronaVac and three doses of CoronaVac^4,14,18,19^. Real-world studies from Hong are Kong and Brazil^9,10^ showed that two doses of CoronaVac provide little protection against mild or moderate COVID-19 diseases associated with Omicron variant across all ages. A homologous booster dose of CoronaVac had no additional protection against symptomatic disease and a moderate increase in protection against severe disease associated with Omicron variant. In this study, we found that a heterologous third dose of Convidecia can effectively elicit Delta and Omicron variant-neutralizing activity in the older immunized individuals, with significantly higher serum-positivity against these variants compared with the three doses of CoronaVac. These findings support the use of a heterologous third Convidecia booster dose could increase the maturity and cross-reactivity of the vaccine-elicited antibodies.

This study has some limitations. Firstly, the number of participants in this study is relatively small. And we did not calculate the sample size for the two-dose regimen, which may lead to an insufficient statistical power. Secondly, only generally healthy old individuals were involved in this study, making it is impossible to evaluate the effects of underling diseases on the immune responses, which might compromise the generalization of the results. Thirdly, we only reported the level of antibody responses collected up to 28 days post-vaccination. Although, the booster doses resulted in a substantial increase in neutralizing antibodies against both wild-type and variants of SARS-CoV-2, quick waning of the antibodies from the peak was also seen after booster doses^20^.

In conclusion, the heterologous prime-boost regimens with the inactivated vaccine CoronaVac and the Ad5-vectored vaccine Convidecia were safe and highly immunogenic in older adults. Heterologous Convidecia booster doses after two doses of CoronaVac markedly improve neutralizing antibody levels against the Delta and Omicron variants. Our findings supported the use of Convidecia as a heterologous booster following the primary immunization with CoronaVac in the old, particularly for countries that primarily use CoronaVac vaccines.

## Author Contributions statement

Pengfei Jin and Jingxin Li had full access to all the data in the study and takes responsibility for the integrity of the data and the accuracy of the data analysis.

### Study concept and design

Fengcai Zhu, Wei Chen, Jingxin Li, Pengfei Jin, Lihua Hou

### Acquisition, analysis, or interpretation of data

Pengfei Jin, Jingxin Li, Xiling Guo, Jiahong Zhu, Fengjuan Shi, Jingxian Liu, Hui Zheng, Yin Chen, Shipo Wu, Xuewen Wang, Xiaoyu Xu

### Drafting of the manuscript

Pengfei Jin, Jingxin Li

### Critical revision of the manuscript for important intellectual content

Fengcai Zhu, Wei Chen

### Statistical analysis

Hui Zheng, Xuewen Wang

### Obtained funding

Fengcai Zhu, Jingxin Li

### Administrative, technical, or material support

Jingxin Li, Pengfei Jin, Xiling Guo, Jinbo Gou, Zhizhou Song, Tao Zhu, Hongxing Pan, Pan Du, Haitao Huang

### Study supervision

Xue Wang, Peng Wan

## Supporting information

Supplement 2_eTables+eFigures

Supplement 1_protocol

CONSORT Check list

## Data Availability

All data produced in the present study are available upon reasonable request to the authors

## Data Availability Statement

The study protocol is available in the Supplementary Information file. Researchers who provide a scientifically sound proposal are allowed to access to the de-identified individual participant data. Individual participant data can be obtained with a request to.

