## Supplement 2_eTables+eFigures for "Heterologous CoronaVac plus Ad5-nCOV versus homologous CoronaVac vaccination among elderly: a phase 4, non-inferiority, randomized study"

### eTable 1. Neutralizing antibodies against SARS-CoV-2 wild-type, and Delta and Omicron variants after boost vaccination.

|  | **Group A (CoronaVac/CoronaVac**  **+Convidecia)** | **Group B (CoronaVac/CoronaVa**  **+CoronaVac)** | ***P* value** | **Group C (CoronaVac**  **+Convidecia)** | **Group D (CoronaVac**  **+CoronaVac)** | ***P* value** |
| --- | --- | --- | --- | --- | --- | --- |
| **Wild-type** | | | | | | |
| Day 14 | | | | | | |
| n/N | 98/98 | 100/100 |  | 47/48 | 46/47 |  |
| GMT | 286.4 | 48.2 | <0.0001 | 70.9 | 9.3 | <0.0001 |
|  | (244.6, 335.2) | (39.5, 58.7) |  | (49.5, 101.7) | (6.2, 13.9) |  |
| Seroconversion  rate (%) | 99.0  (94.5,99.8) | 98  (93.0, 99.5) | 0.5667 | 93.6  (82.8, 97.8) | 58.7  (44.3, 71.7) | <0.0001 |
| GMFI | 143.2  (122.3, 167.6) | 24.1  (19.8, 29.4) | <0.0001 | 35.5  (24.8, 50.8) | 4.6  (3.1, 6.9) | <0.0001 |
| GMT ratio | 6.2  (4.7, 8.1) | |  | 7.6  (4.1,14.1) | |  |
| Day 28 | | | | | | |
| n/N | 97/97 | 100/100 |  | 47/47 | 44/44 |  |
| GMT | 243.5  (209.7, 282.9) | 42.2  (34.4, 51.8) | <0.0001 | 51.3  (38.3, 68.6) | 7.2  (5.0, 10.3) | <0.0001 |
| Seroconversion  rate (%) | 100.0  (96.2, 100.0) | 98.0  (93.0, 99.5) | 0.1615 | 100.0  (92.4, 100.0) | 52.3  (37.9, 66.2) | <0.0001 |
| GMFI | 121.8  (104.8, 141.4) | 21.1  (17.2, 25.9) | <0.0001 | 25.7  (19.2, 34.3) | 3.6  (2.5, 5.2) | <0.0001 |
| GMT ratio | 5.8  (4.3, 7.7) | |  | 7.2  (4.2,12.1) | |  |
| **Delta, B.1.617.2** | | | | | | |
| Day 14 | | | | | | |
| n/N | 50/50 | 50/50 |  | 24/30 | 23/30 |  |
| GMT | 45.9  (34.5, 61.0) | 7.3  (5.6, 9.5) | <0.0001 | 5.0  (3.2, 7.9) | 4.0  (2.4, 6.7) | 0.4876 |
| Seropositive  rate (%) | 100.0  (92.9, 100.0) | 80.0  (67.0, 88.8) | 0.4438 | 58.3  (38.8, 75.5) | 30.4  (15.6, 50.9) | 0.2312 |
| GMT ratio | 6.3  (3.4, 12.4) | |  | 1.3  (1.0, 2.7) | |  |
| **Omicron, BA.1.1** | | | | | | |
| Day 14 | | | | | | |
| n/N | 50/50 | 50/50 |  | 24/30 | 23/30 |  |
| GMT | 32.9  (23.1, 46.9) | 4.4  (3.2, 5.9) | <0.0001 | 4.8  (2.8, 8.2) | 3.7  (2.2, 6.1) | 0.4707 |
| Seropositive  rate (%) | 94.0  (83.8, 97.9) | 42.0  (29.4, 55.8) | 0.0138 | 45.8  (27.9, 64.9) | 30.4  (15.6, 50.9) | 0.4672 |
| GMT ratio | 7.5  (3.5, 18.0) | |  | 1.3  (1.0, 2.9) | |  |

Data shown are the geometric mean (95% CI) for continuous variables and the percent (95% CI) for binary variables. N= the number of participants included in the per-protocol cohort, n=the number of samples analyzed. Seroconversion was defined as at least a fourfold increase in the antibody titers after boost immunization compared to the baseline level. GMT Ratio was defined as the ratio of GMT of neutralizing antibodies elicited by the heterologous arm compared to the corresponding homologous arm. Seropositive was defined as the antibody titers≥4. GMT=geometric mean titer; GMFI=geometric mean fold increase.

### eTable 2. Anti-receptor binding domain (RBD) IgG antibody levels before and after boost vaccination.

|  | **Group A (CoronaVac/CoronaVac+Convidecia)** | **Group B (CoronaVac/CoronaVa+**  **CoronaVac)** | ***P* value** | **Group C (CoronaVac+**  **Convidecia)** | **Group D (CoronaVac+**  **CoronaVac)** | ***P* value** |
| --- | --- | --- | --- | --- | --- | --- |
| Day 0 | | | | | | |
| n/N | 99/99 | 100/100 |  | 50/50 | 50/50 |  |
| GMC  (BAU /ml) | 22.6 | 23.4 | 0.7887 | 11.5 | 11.9 | 0.7374 |
|  | (19.1, 26.7) | (19.2, 28.5) |  | (10.3, 12.9) | (10.2, 13.8) |  |
| Day 14 | | | | | | |
| n/N | 98/98 | 100/100 |  | 48/48 | 47/47 |  |
| GMC (BAU/ml) | 1731.9 | 253.8 | <0.0001 | 724.0 | 94.7 | <0.0001 |
|  | (1451.6, 2066.4) | (214.7,300.1) |  | (477.0, 1098.8) | (60.7, 147.6) |  |
| Seroconversion | 100 | 79 | <0.0001 | 95.7 | 69.8 | 0.001 |
| rate (%) | (96.2, 100.0) | (70.0, 85.8) |  | (85.8, 98.8) | (54.9, 81.4) |  |
| GMFI | 76.7 | 10.9 | <0.0001 | 63.0 | 8.0 | <0.0001 |
|  | (63.6, 92.6) | (8.5, 13.8) |  | (41.9, 94.8) | (5.3, 11.9) |  |
| Day 28 | | | | | | |
| n/N | 97/97 | 100/100 |  | 47/47 | 44/44 |  |
| GMC (BAU/ml) | 1355.9 | 227 | <0.0001 | 653.9 | 91.5 | <0.0001 |
|  | (1141.4, 1610.7) | (190.2, 270.9) |  | (465.9, 917.9) | (61.6, 136.0) |  |
| Seroconversion rate (%) | 100  (96.2, 100.0) | 76  (66.8, 83.3) | <0.0001 | 97.9  (88.9, 99.6) | 74.4  (59.8, 85.1) | 0.0011 |
| GMFI | 60.1  (49.6, 72.7) | 9.7  (7.7, 12.2) | <0.0001 | 56.9  (40.7, 79.6) | 7.7  (5.4, 11.0) | <0.0001 |

Data shown are the geometric mean (95% CI) for continuous variables and the percent (95% CI) for binary variables. N= the number of participants included in the modified per-protocol cohort, n=the number of samples analyzed. Seroconversion was defined as at least a fourfold increase in the antibody titers after boost immunization compared to the baseline level. GMC=geometric mean concentration; GMFI=geometric mean fold increase.

### eTable 3. Solicited and unsolicited adverse reactions that occurred within 28 days after vaccination

|  |  | **Group A (CoronaVac/CoronaVac**  **+Convidecia, n=99)** | **Group B (CoronaVac/CoronaVa**  **+CoronaVac, n=100)** | ***P***  **value** | **Group C (CoronaVac+Convidecia, n=50)** | **Group D (CoronaVac+CoronaVac, n=50)** | ***P***  **value** |
| --- | --- | --- | --- | --- | --- | --- | --- |
| **Solicited adverse reactions within 28 days** | | | | | | | |
| Total | Any | 8 (8.1) | 4 (4.0) | 0.227 | 8 (16.0) | 1 (2.0) | 0.031 |
|  | Severe | 0 | 0 | - | 1 (2.0) | 0 | >0.999 |
| **Injection site adverse reactions** | | | | | | | |
| Total | Any | 4 (4.0) | 3 (3.0) | 0.721 | 6 (12.0) | 1 (2.0) | 0.112 |
|  | Severe | 0 | 0 | - | 1 (2.0) | 0 | >0.999 |
| Pain | Any | 2 (2.0) | 3 (3.0) | >0.999 | 6 (12.0) | 1 (2.0) | 0.112 |
|  | Severe | 0 | 0 | - | 1 (2.0) | 0 | >0.999 |
| Induration | Any | 1 (1.0) | 0 | 0.497 | 2 (4.0) | 0 | 0.495 |
| Swelling | Any | 1 (1.0) | 0 | 0.497 | 2 (4.0) | 0 | 0.495 |
| Itching | Any | 1 (1.0) | 0 | 0.497 | 2 (4.0) | 0 | 0.495 |
| Erythema | Any | 0 | 0 | - | 2 (4.0) | 0 | 0.495 |
| **Systemic adverse reactions** | | | | | | | |
| Total | Any | 4 (4.0) | 1 (1.0) | 0.212 | 2 (4.0) | 0 | 0.495 |
| Fever | Any | 2 (2.0) | 1 (1.0) | 0.621 | 1 (2.0) | 0 | >0.999 |
| Fatigue | Any | 2 (2.0) | 0 | 0.246 | 2 (4.0) | 0 | 0.495 |
| Myalgia | Any | 1 (1.0) | 1 (1.0) | >0.999 | 0 | 0 | - |
| Headaches | Any | 1 (1.0) | 0 | 0.497 | 0 | 0 | - |
| Fainting | Any | 1 (1.0) | 0 | 0.497 | 0 | 0 | - |
| Joint pain | Any | 0 | 0 | - | 1 (2.0) | 0 | >0.999 |
| Diarrhea | Any | 0 | 0 | - | 1 (2.0) | 0 | >0.999 |
| **Unsolicited adverse reactions within 28 days** | | | | | | | |
| Dizziness | Any | 1 (1.0) | 0 | 0.497 | 0 | 0 | - |

Data are n (%). n = number of participants. % = proportion of participants. Any = all the participants with any grade adverse reactions or events.

### eTable 4. List of severe adverse events

| **Group** | **Disease** | **Time of vaccination** | **Start time** | **End time** | **Outcome** | **Correlation with vaccination** |
| --- | --- | --- | --- | --- | --- | --- |
| Group C | Internal rheumatoid arthritis | 26 August, 2021 | 12 February, 2022 | 18 February, 2022 | Improvement | NO |
| Group D | [Chronic](javascript:;) [bronchitis](javascript:;) with emphysema | 26 August, 2021 | 10 January, 2022 | 18 January, 2022 | Improvement | NO |
| Group D | [Chronic](javascript:;) [bronchitis](javascript:;) with emphysema | 26 August, 2021 | 19 December, 2021 | 26 December, 2021 | Improvement | NO |
| Group A | Acute inferior posterior wall myocardial infarction | 11 November, 2021 | 28 December, 2021 | 4 January, 2022 | Improvement | NO |

### eTable 5. Multivariate linear regression analysis of age and neutralizing antibody titers 14 days after boost vaccination.

| **Strain variable** | **Independent variable** | **β** | **Significance** | **VIF** | **R^2^** |
| --- | --- | --- | --- | --- | --- |
| **Three-dose regimen cohorts** | | | | | |
| Neutralizing antibodies against wild-type SARS-CoV-2 | Constant | -23.019 | 0.812 |  | 0.452 |
|  | Group | **224.898** | **0.000** | 1.185 |  |
|  | Time since the last priming dose of inactivated vaccine (months) | 63.640 | 0.270 | 1.330 |  |
|  | Age | -2.823 | 0.931 | 1.128 |  |
|  | Gender | 63.820 | 0.273 | 1.377 |  |
|  | Neutralizing antibodies against wild-type SARS-CoV-2 before boosting | -0.394 | 0.981 | 1.400 |  |
| Neutralizing antibodies against Delta variant | Constant | 25.532 | 0.434 |  | 0.534 |
|  | Group | **46.683** | **0.014** | 1.140 |  |
|  | Time since the last priming dose of inactivated vaccine (months) | -20.717 | 0.295 | 1.329 |  |
|  | Age | -20.570 | 0.073 | 1.116 |  |
|  | Gender | 38.208 | 0.052 | 1.260 |  |
|  | Neutralizing antibodies against Delta variant before boosting | 13.140 | 0.001 | 1.259 |  |
| Neutralizing antibodies against Omicron variant | Constant | 11.788 | 0.490 |  | 0.402 |
|  | Group | **27.541** | **0.002** | 1.062 |  |
|  | Time since the last priming dose of inactivated vaccine (months) | 10.526 | 0.216 | 1.168 |  |
|  | Age | -6.095 | 0.313 | 1.172 |  |
|  | Gender | 11.992 | 0.165 | 1.192 |  |
| **Two-dose regimen cohorts** | | | | | |
| Neutralizing antibodies against wild-type SARS-CoV-2 | Constant | 43.990 | 0.304 |  | 0.162 |
|  | Group | **99.116** | **0.000** | 1.007 |  |
|  | Age | -4.289 | 0.426 | 1.030 |  |
|  | Gender | 3.369 | 0.839 | 1.004 |  |
|  | Neutralizing antibodies against wild-type SARS-CoV-2 before boosting | -4.868 | 0.766 | 1.032 |  |
| Neutralizing antibodies against Delta variant | Constant | 43.770 | 0.434 |  | 0.118 |
|  | Group | **31.527** | **0.005** | 1.018 |  |
|  | Age | -11.799 | 0.036 | 1.012 |  |
|  | Gender | 10.137 | 0.357 | 1.023 |  |
|  | Neutralizing antibodies against Delta variant before boosting | -11.459 | 0.675 | 1.019 |  |
| Neutralizing antibodies against Omicron variant | Constant | 100.766 | 0.016 |  | 0.110 |
|  | Group | 5.205 | 0.554 | 1.116 |  |
|  | Age | -19.130 | 0.032 | 1.009 |  |
|  | Gender | -3.114 | 0.724 | 1.120 |  |

The independent variables are group (For three-dose regimen, group B as reference; For two-dose regimen, group D as reference), age (categorica variable, 18-29 years as reference, 30-39 years, 40-49 years, 50-59 years, 60-69 years, and ≥70 years), gender (female as reference), time since the last priming dose of inactivated vaccine (months) (categorica variable, ≤4 months as reference and >4 months) and log-transformed neutralizing antibody titers before boosting (continuous variable) included in a multivariable analysis.

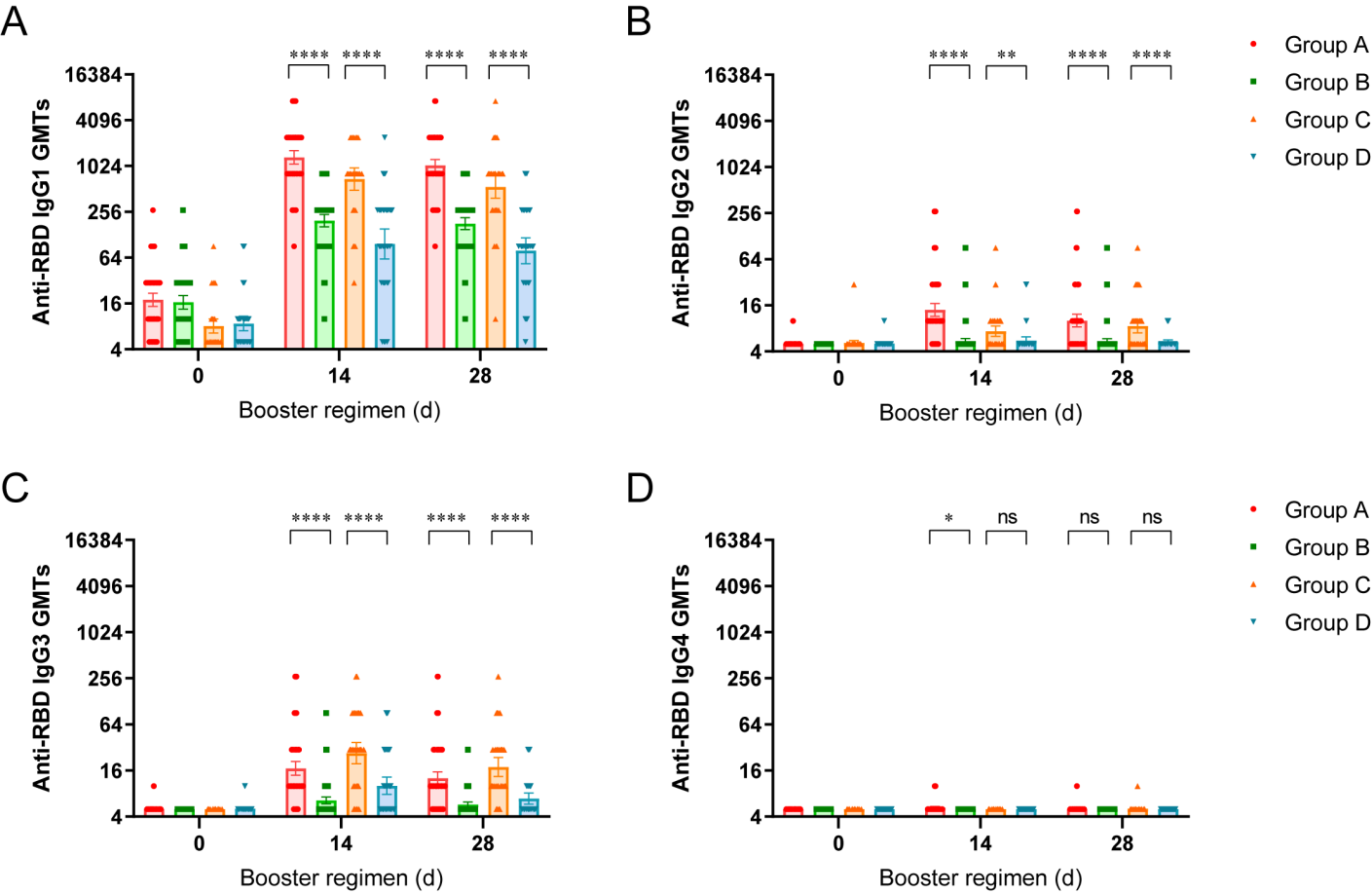

### eFigure 1. Anti-receptor binding domain (RBD) binding IgG isotypes before and after boost vaccination

(A) GMTs of anti-RBD IgG1 isotypes, (B) IgG2 isotypes, (C) IgG3 isotypes, and (D) IgG4 isotypes. Data are GMT (95% CI). Error bars indicate 95% CIs. Group A, primed with two doses of CoronaVac and given one dose of Convidecia (n= 98); Group B, primed with two doses of CoronaVac and given one dose of CoronaVac (n= 100); Group C, primed with one dose of CoronaVac and given one dose of Convidecia (n= 48); Group D, primed with one dose of CoronaVac and given one dose of CoronaVac (n= 47). All the paired data of RBD-binding antibodies from participants are included in the analysis. The p values are the results of a comparison between the two treatment groups using a T-test for log-transferred antibody titers (Group A vs. Group B, and Group C vs. Group D). The discrepancies between the numbers of data points presented in the figures and the numbers of participants in the groups are due to the overlapping of the dots. *P-value <0.05; **P-value <0.01; ****P value <0.0001; ns, representing P>0.05. GMT=geometric mean titer.

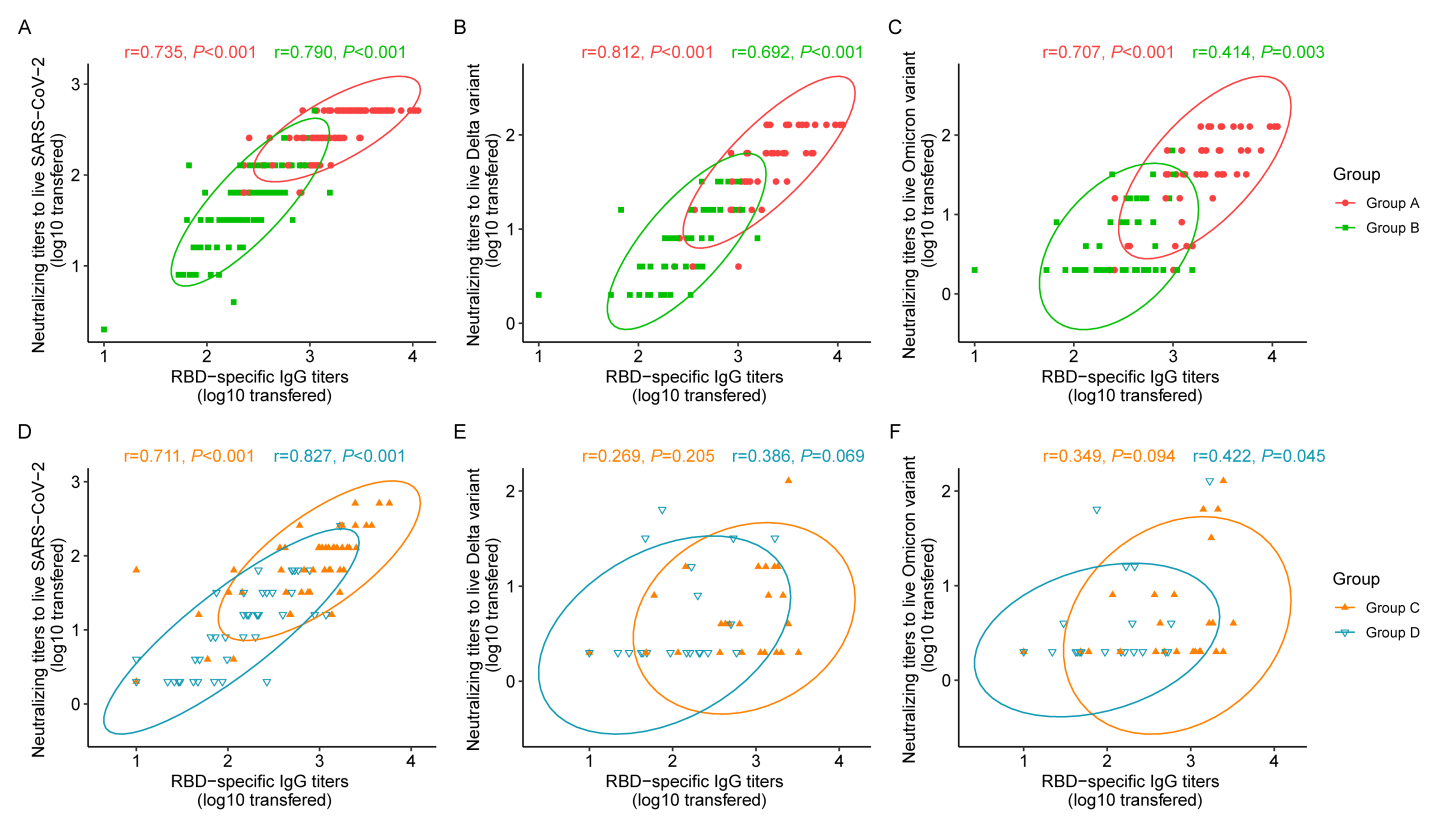

### eFigure 2. Correlation between anti-receptor binding domain (RBD) binding IgG and neutralizing antibody titers

Correlation of RBD-specific antibodies with neutralizing antibodies against wild-type SARS-CoV-2 (A, D), Delta variant (B, E), and Omicron variant (C, F) 14 days after boosting, respectively. Pearson correlation coefficients (95% CIs) and P values are presented for each group. The ellipses show the 95% CIs by assuming a multivariate normal distribution. Group A, primed with two doses of CoronaVac and given one dose of Convidecia; Group B, primed with two doses of CoronaVac and given one dose of CoronaVac; Group C, primed with one dose of CoronaVac and given one dose of Convidecia; Group D, primed with one dose of CoronaVac and given one dose of CoronaVac. All the paired data of neutralizing antibodies and RBD-binding antibodies from participants are included in the analysis. The discrepancies between the numbers of data points presented in the figures and the numbers of participants in the groups are due to the overlapping of the dots.

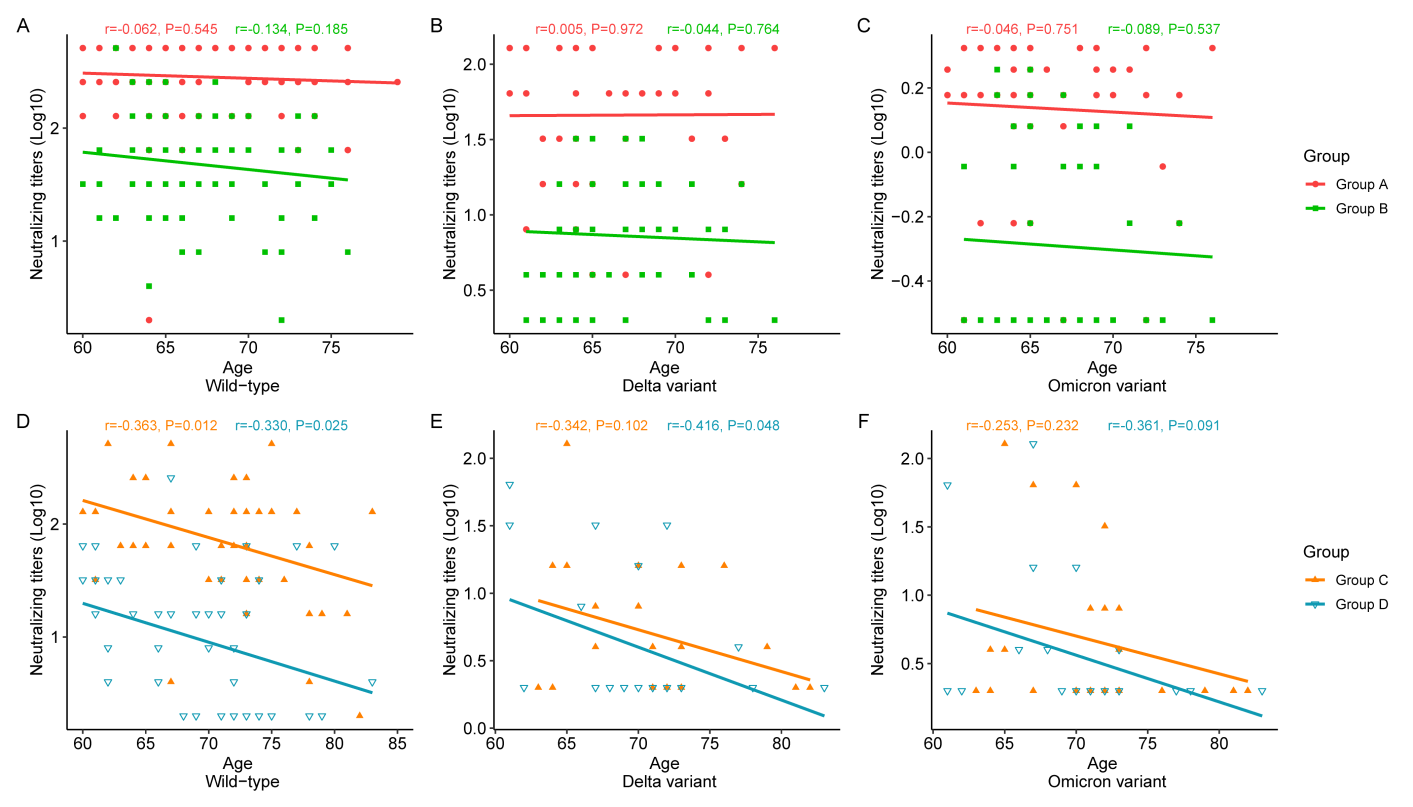

### eFigure 3. **Correlation of age and neutralizing antibodies titers 14 days after boost vaccination**

Correlation of age and neutralizing antibodies titers to SARS-CoV-2 wild-type (A, D), Delta variant (B, E) and Omicron variant (C, F) 14 days after boosting by four immunization regimens. Pearson correlation coefficients and P values are presented for each group. Group A, primed with two doses of CoronaVac and given one dose of Convidecia; Group B, primed with two doses of CoronaVac and given one dose of CoronaVac; Group C, primed with one dose of CoronaVac and given one dose of Convidecia; Group D, primed with one dose of CoronaVac and given one dose of CoronaVac.

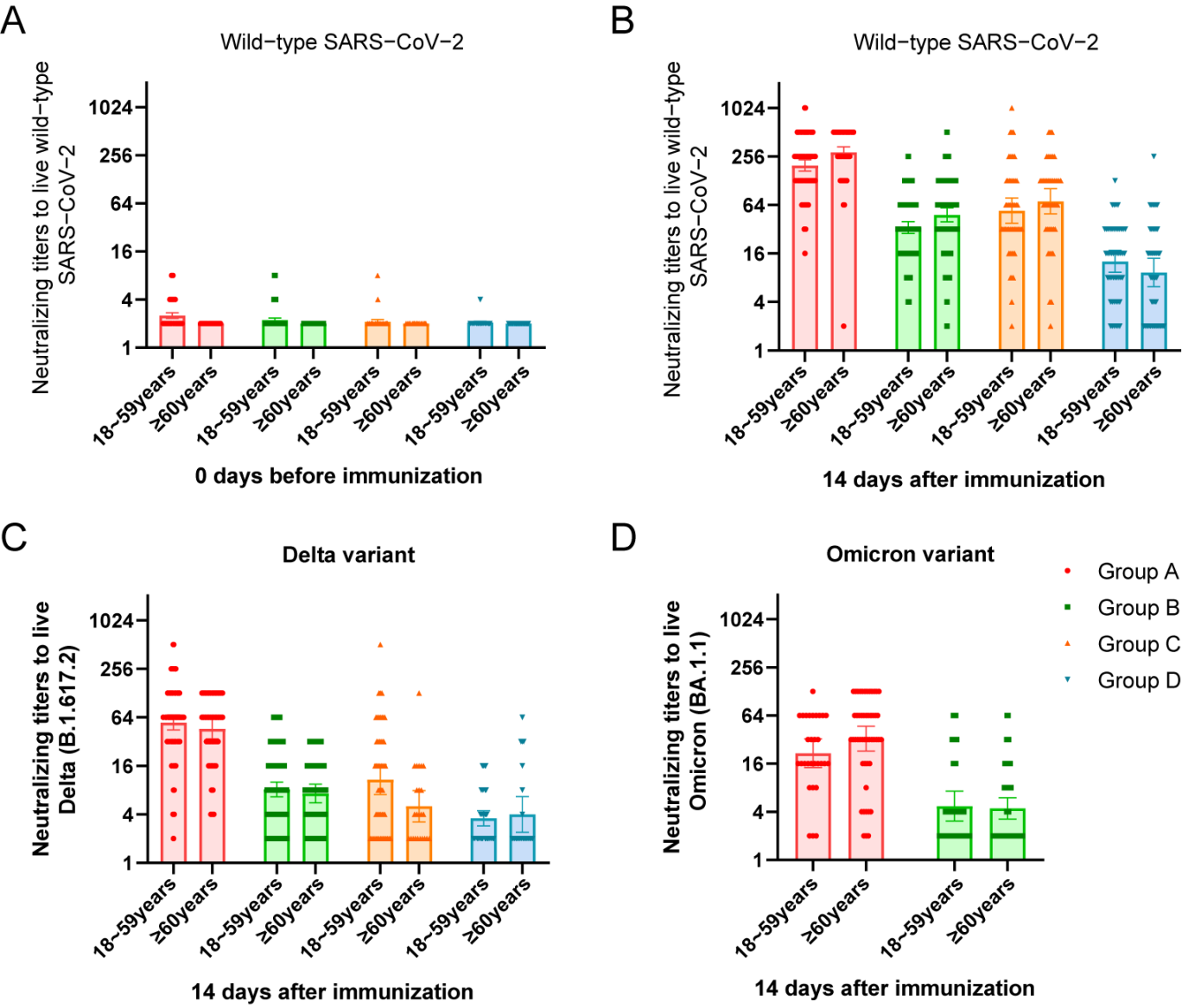

### eFigure 4. GMTs of neutralizing antibody titers before and after boost vaccination according to age.

GMTs of neutralizing antibodies against SARS-CoV-2 wild-type before boosting (A), and neutralizing antibodies against wild-type strain (B) and Delta variant (C) and Omicron variant (D)14 days after boosting by age group (18-60 years and ≥60 years) and immunization regimens. Group A, primed with two doses of CoronaVac and given one dose of Convidecia; Group B, primed with two doses of CoronaVac and given one dose of CoronaVac; Group C, primed with one dose of CoronaVac and given one dose of Convidecia; Group D, primed with one dose of CoronaVac and given one dose of CoronaVac. GMT=geometric mean titer.
