## Supplement 1_protocol for "Heterologous CoronaVac plus Ad5-nCOV versus homologous CoronaVac vaccination among elderly: a phase 4, non-inferiority, randomized study"

**Study on heterologous prime-boost immunization of Inactivated SARS-CoV-2 vaccine (Vero cells) and Recombinant COVID-19 vaccine (Ad5 Vector) in healthy adults aged 60 years and older**

Project number: JSVCT117

Principal Investigator: Jingxin Li

Research unit: Jiangsu Provincial Center for Disease

Control and Prevention

Version number: Version 1.0

Version date: April 16, 2021

| Research Title | Study on heterologous prime-boost immunization of Inactivated SARS-CoV-2 vaccine (Vero cells) and Recombinant COVID-19 vaccine (Ad5 Vector) in healthy adults aged 60 years and older | |
| --- | --- | --- |
| Detailed description of the topic | Safety and immunogenicity of heterologous prime-boost immunization of Inactivated SARS-CoV-2 vaccine (Vero cells) and Recombinant COVID-19 vaccine (Ad5 Vector) in healthy adults aged 60 years and older in China: a randomized, observer-blind, parallel-controlled clinical study | |
| Item No. | JSVCT117 | |
| Research | Jiangsu Provincial Center for Disease Control and Prevention | |
| Research Vaccines | Novel inactivated coronavirus vaccine (Vero cells), recombinant novel coronavirus vaccine (type 5 adenovirus vector) | |
| Program Date | April 16, 2021 | |
| Version number | Version 1.0 | |
| Principal Investigator | Li Jingxin | Chief Physician  Jiangsu Provincial Center for Disease Control and Prevention |
| Authors: Li Jingxin, Jin Pengfei, Zhu Fengcai  Jiangsu Provincial Center for Disease Control and Prevention  Copyright © 2021 Unauthorized Reproduction or Use Prohibited | | |

**Principal Investigator Statement**

I agree.

- Assume principal investigator responsibilities for this clinical study.
- Ensure that this study is conducted following the study protocol and the field standard operating manual.
- Ensure that no changes are made to the trial protocol without review and written approval by the ethics committee, except when required to eliminate immediate harm to the subject or to comply with the requirements of the pharmaceutical administration (e.g., aspects involving administration).
- I fully grasp the proper use of the study vaccine as described in the protocol.
- I am familiar with and will comply with the Good Clinical Practice (GCP) and all relevant regulatory requirements for the management of drug clinical trials.

| Research Title | Study on heterologous prime-boost immunization of Inactivated SARS-CoV-2 vaccine (Vero cells) and Recombinant COVID-19 vaccine (Ad5 Vector) in healthy adults aged 60 years and older | |
| --- | --- | --- |
| Detailed description of the research topic | Safety and immunogenicity of heterologous prime-boost immunization of Inactivated SARS-CoV-2 vaccine (Vero cells) and Recombinant COVID-19 vaccine (Ad5 Vector) in healthy adults aged 60 years and older in China: a randomized, observer-blind, parallel-controlled clinical study | |
| Research Vaccines | Novel inactivated coronavirus vaccine (Vero cells), recombinant novel coronavirus vaccine (type 5 adenovirus vector) | |
| Item No. | JSVCT117 | |
| Program Date | April 162021 | |
| Version number | Version 1.0 | |
| Principal Investigator | Name：Jingxin Li  Title: Chief Physician  Position: Deputy Director, Institute of Clinical Evaluation of Vaccines, Jiangsu Provincial Center for Disease Control and Prevention  Unit: Jiangsu Provincial Center for Disease Control and Prevention  Address: Room 306, Building B, No. 172 Jiangsu Road, Nanjing  Zip code: 210009  Phone.18915999772  Fax: 025-83759529  | |
| Principal Investigator (Signature) |  | Date of signature. |

**Program revision record**

| Serial number | Revision section | Revision Notes | Reason for revision |
| --- | --- | --- | --- |

**Abbreviations**

| **Term/ Abbreviation** | **Definition/ Full Form** |
| --- | --- |
| AE | Adverse Events |
| AR | Adverse reactions |
| Ad5 | Replication-deficient human5 adenovirus |
| CDC | Centers for Disease Control and Prevention |
| COVID-19 | Corona Virus Disease 2019 |
| eCRF | Electronic Case Report Form |
| ELISA | Enzyme-linked immunosorbent assay |
| FAS | Full analysis set |
| GCP | Quality management standard for drug clinical trials |
| GMI | Geometric Mean Fold Increase |
| GMT | Geometric mean titer |
| IEC | Independent Ethics Committee |
| ITT | Intent-to-treat |
| NIFDC | National Institute for Food and Drug Control |
| NMPA | National Medical Products Administration |
| PPS | Per Protocol Set |
| SAE | Serious Adverse Events |
| SARS-CoV-2 | Severe Acute Respiratory Syndrome Coronavirus 2 |
| SS | Safety Set |
| VP | Virus particles |

Catalog

Program Summary

| Research Title | Study on heterologous prime-boost immunization of Inactivated SARS-CoV-2 vaccine (Vero cells) and Recombinant COVID-19 vaccine (Ad5 Vector) |
| --- | --- |
| Detailed description of the research topic | Safety and immunogenicity of heterologous prime-boost immunization of Inactivated SARS-CoV-2 vaccine (Vero cells) and Recombinant COVID-19 vaccine (Ad5 Vector) in healthy adults aged 60 years and older in China: a randomized, observer-blind, parallel-controlled clinical study |
| Disease Prevention | Prevention of COVID-19 caused by SARS-CoV-2 infection |
| Study population | healthy adults aged 60 years and older |
| Number of Subjects | About 300 subjects |
| Purpose of the study | To evaluate the safety and immunogenicity of a heterologous  prime-boost immunization of Inactivated SARS-CoV-2 vaccine  (Vero cells) and Recombinant COVID-19 vaccine (Ad5 Vector) in healthy adults aged 60 years and older. |
| Research site | Lianshui County Center for Disease Control and Prevention |
| Fundamentals | Vaccines are one of the most effective ways to control the COVID-19 global pandemic. Currently, there are five major research and development technology routes of COVID-19 vaccine worldwide, namely, inactivated vaccine, viral vectored vaccine, live attenuated vaccine, recombinant protein vaccine, and nucleic acid vaccine. The inactivated COVID-19 vaccine is developed by Beijing Sinovac Research & Development Co., Ltd., and the COVID-19 vaccine (human adenovirus 5 vector) is developed by the Military Academy of Military Medical Institute and CanSino Biologics Inc. These vaccines have got conditional approval on the market in China. Foreign phase 3 preliminary clinical trials showed that the short-term efficacy of COVID-19 against symptomatic COVID-19 was between 50% and 70%. Both vaccines have met the requirement of WHO for the minimum efficacy of 50%, however, the efficacy was moderate. Compared to the 95% efficacy of the mRNA vaccine against COVID-19 developed by Moderna and Pfizer/ Biotech, the inactive vaccine and Ad5 vectored vaccine seemed to have lower efficacy.  To induce a sufficiently high level of immune response, Russia took the lead in the world in forming an immunization program with two different viral vector-based vaccines-- rAd26/rAd5 vectored vaccine. The reported protection efficacy of the rAd26/rAd5 vectored vaccine was 91.4%. The United Kingdom has also announced a study sequential vaccination of adenovirus vectored COVID-19 vaccine and mRNA vaccine, to optimize the immunization program of the existing COVID-19 vaccine and achieve better protective effect in a short time.  In theory, the types and characteristics of immune responses induced by the Ad5 vectored vaccine and inactivated vaccine are different, and the sequential vaccination of the two vaccines at different time points may improve the quality of the immune response, and optimize the existing immunization strategies. |
| Test vaccines | Investigational vaccine 1: Inactivated COVID-19 vaccine (Vero cells)  Manufacturer: Beijing Institute of Biological Products/SINOVAC BIOTECH CO., LTD.  Specification: 0.5ml/bottle, for commercial packaging, storage  according to the instructions.    Investigational vaccine 2: Recombinant COVID-19 vaccine (Ad5 Vector)  Manufacturer: Academy of Military Medicine, Academy of  Military Sciences/CanSino Biologics Inc.  Specification: 0.5ml/bottle, for commercial packaging, storage  according to the instructions.  Immunization procedures:  Four study groups were included in this study: heterologous booster immunization group (group A), conventional booster immunization group (group B), heterologous immunization group (group C), and conventional immunization group (group D).  Recruited subjects in groups A and B completed 2 doses of Inactivated COVID-19 vaccine (Vero cells) basic immunization according to the 0-28 days immunization program. Subjects in group A will receive 1 dose of Recombinant COVID-19 vaccine (Ad5 Vector) from 3 - 6 months after 2 doses of Inactivated COVID-19 vaccine (Vero cells) basic immunization. Subjects in group B will receive 1 dose of Inactivated COVID-19 vaccine (Vero cells) from 3 - 6 months based on 2 injections of the Inactivated COVID-19 vaccine (Vero cells).  Recruited subjects in groups C and D have been immunized with 1 dose of inactivated COVID-19 vaccine. In group C, subjects will be immunized with 1 dose of recombinant COVID-19 vaccine (Ad5 vector) from 1-3 months based on 1 dose of inactivated COVID-19 vaccine. In group D, the subjects will receive another dose of inactivated COVID-19 vaccine from 1-3 months based on one injection of the inactivated COVID-19 vaccine.  Immunization dosages:  Inactivated COVID-19 vaccine (Vero cells): 0.5 ml for a single injection.  Recombinant COVID-19 vaccine (Ad5 vector) : single injection 0.5ml, containing 5.0×10VP^10^.  Immunization route:  The site of inoculation is the lateral deltoid muscle of the upper arm, and the route of inoculation is an intramuscular injection.  Storage and transport conditions:  Store and transport at 2~8℃ away from light and strictly prevent from freezing. |
| Trial design | Study design: Single-center, randomized, observer-blinded, parallel-controlled sequential immune clinical trial. Calculation of sample size: （1） Hypothesis 1：GMT of Group A is not inferior to that in group B at Day28 after the boost vaccination. （2） Hypothesis 2: GMT of Group A is superior to that in group B at Day28 after the boost vaccination. *If hypothesis 1 holds, further statistical inferences can be made about hypothesis 2. *The comparison of GMT after immunization in group C and group D was used as the initial exploration content, and sample size estimation without statistical inference was done. Considering the recruitment difficulty of the population of Group C and Group D, it was set to half of the sample size of Group A and Group B. Three to six months after two doses of inactivated vaccine, the baseline GMT level before the booster is expected to be about 1:40 (log10X=1.6), and 1:80 (log10X =1.9) after one dose of inactivated vaccine. After receiving one dose of the recombinant COVID-19 vaccine (Ad5 vector) as a booster, the GMT is estimated to reach 1:160 (log10X =2.2). The standard deviation is about 40 (log10X=0.6), the sample size is calculated: Hypothesis 1, one-sided α = 0.025 significance level, 90% study power, GMT ratio of group A/group B Non-inferiority Margin is 0.67 (log10X =-0.174), the ratio of group A and group B is 1:1, and the sample size is 35 per group. Hypothesis 2, one-sided α = 0.025 significance level, and the ratio of group A and group B are 1:1. To ensure a 90% study power, a sample size of 98 per group could show that the GMT level of group A after immunization is better than that of group B. Therefore, two hypotheses were considered to be satisfied, with a sample size of approximately 100 individuals each in the sequential booster immunization group (Group A) and the conventional booster immunization group (Group B), and approximately 50 individuals in the sequential immunization group (Group C) and the conventional immunization group (Group D). The total sample size is approximately 300 individuals. Table 1. sample size of each study group Code \| Grouping \| Sample size \| Basic immunity \| Vaccines \| Sequential immunization; A \| Sequential booster immunization group \| 100 \| Completion of two doses of inactivated vaccine \| Adenovirus vector vaccine (1 dose) \| Sequential immunization after the second dose of inactivated vaccine 3~6 months; B \| Routine booster immunization group \| 100 \| \| Inactivated vaccine (1 dose) \|; C \| Sequential immunization group \| 50 \| Completion of one dose of inactivated vaccine \| Adenovirus vector vaccine (1 dose) \| Sequential immunization 1~2 months after the first dose of inactivated vaccine; D \| Conventional immunization group \| 50 \| \| Inactivated vaccine (1 dose) \|; \| \| 300 \| \| \| Note: Subjects in Groups A and B must have completed 2 doses of inactivated vaccine immunization. Subjects in Group C and Group D must have completed 1 dose of inactivated vaccine immunization. Randomization and Blinding: Eligible subjects are stratified into groups that have completed 2 doses of basic immunization (groups A and B) and groups that have completed 1 dose of basic immunization (groups C and D). By using the method of block randomization, the subjects are randomly assigned to sequential booster immunization group (Group A), conventional booster immunization group (Group B), sequential immunization group (Group C), and conventional immunization group (Group D) in a ratio of 2:2:1:1. The subjects' randomization table is generated by an independent randomization professional using SAS version 9.4 or above and imported into the Interactive Response Technology (IRT) system. The allocation of the treatment groups is accessible only to authorized unblinding staff. Subjects, investigators, and the sponsor's research management team will be blinded throughout the trial. The unblinding staff at authorized research centers can obtain grouping information of subjects through the IRT system and use the investigational vaccine for the corresponding group based on the grouping information. This study was conducted by setting up unblinded staff to dispense, configure, and administer the vaccine. Non-blinded personnel was not involved in field work other than dispensing, configuration, and vaccination, and all other study staff remained blinded. Study plan: This project plans to recruit 300 healthy subjects aged 60 years and older. Subjects who gave informed consent and passed the interrogation screening were randomly assigned to sequential booster immunization group (Group A) or routine booster immunization group (Group B), and sequential immunization group (Group C) or routine immunization group (Group D). given a booster dose of inactivated COVID-19 vaccine or recombinant COVID-19 vaccine (Ad5 vector) at 3 to 6 months after the completion of the basic immunization of 2 doses of inactivated COVID-19 vaccine. The volunteers in groups C and D are required to complete the basic immunization of 1 injection of inactivated COVID-19 vaccine, and after the 1-2 months enrolled subjects are given the second injection with inactivated COVID-19 vaccine or recombinant COVID-19 vaccine (Ad5 vector) based on one injection of inactivated COVID-19 vaccine. All enrolled subjects collected blood samples at day 0 (before vaccination), and at day14, day28, and 6 months after the boost vaccination to detect serum antibody level or cellular immune response level, respectively. The four groups are enrolled in parallel. During the enrollment process, safety data are evaluated in real-time. Once safety problems (suspension or termination criteria) of vaccination are found, late enrollment will be immediately suspended or terminated. Study duration: Each subject will remain in this study for approximately 6 months from enrollment to discharge from the last visit. |
| Endpoint Indicators | Primary study endpoint:   - Incidence of adverse reactions within 28 days after sequential immunization in each group. - Neutralizing antibodies to live SARS-CoV-2 (GMT) after sequential immunization 14 days in each group.   Secondary study endpoints:  1. Security endpoint   - Incidence of recruiting adverse events on day 0-14 after sequential immunization in each group. - Incidence of non-recruiting adverse events on day 0-28 after sequential immunization in each group. - Incidence of serious adverse events within 6 months after sequential immunization in each group.   2. Humoral immunity study endpoints   - Geometric mean titers (GMT) of anti-SARS-CoV-2 S and N protein-specific antibody (ELISA) at Day14, Day28, and Month 6 after sequential immunization in each group. - GMT of neutralizing antibodies to live SARS-CoV-2 at day 28 and months 6 after sequential immunization in each group. - The Geometric Mean of the Fold Increase (GMFI) of antibody level of anti-SARS-CoV-2 S and N protein-specific antibody (ELISA) at Day14, Day28 and Month 6 after sequential immunization compared with Day0 before sequential immunization in each group - The Geometric Mean of the Fold Increase (GMFI) of neutralizing antibodies to live SARS-CoV-2 at Day14, Day28, and Month 6 after sequential immunization compares with Day0 before sequential immunization in each group. - The positive conversion rate of antibody level of anti-SARS-CoV-2 S and N protein-specific antibody (ELISA) at Day14, Day28, and Month 6 after sequential immunization compared with Day0 before sequential immunization in each group (≥ 4-fold increase). - Positive conversion rates of neutralizing antibodies to live SARS-CoV-2 at Day14, Day28, and Month 6 after sequential immunization compare with Day0 before sequential immunization in each group (≥4-fold increase).  3. Cellular immunology study endpoints  - Levels of IFN-γ, TNF-α, IL-5, IL-4, and IL-13 were secreted by specific T cells at Day14 after sequential exemption in each group.   Exploratory study endpoints.   - Types of IgG antibodies against SARS-CoV-2 S protein at Day14, Day28, and Month 6 after sequential immunization in each group. - Cross-neutralization of anti-SARS-CoV-2 neutralizing antibodies produced after sequential immunization of each group with other coronaviruses. - Differentiation of major immune cell populations such as B cells and T cells and antibody profiles at Day14, Day28, and Month 6 after sequential immunization in each group. |
| Visiting Plan | This study requires to complete four visits, respectively:  V1: Day 0 (sequential immunization), informed consent and signature, physical examination, questioning and screening, pre-immunization blood collection, sequential immunization, and observation to be completed.  V2: Day 14 after sequential immunization, a safety follow-up visit, and blood collection need to be completed.  V3: On day 28 after sequential immunization, a safety follow-up visit with blood collection needs to be completed.  V4: On Month 6 after sequential immunization, a safety follow-up and blood collection should be completed |
| Criteria for  pausing or early  termination | Criteria for pausing:  - Occurrence of one or more ≥grade 4 adverse reaction or serious adverse event that may be associated with vaccination.  - Occurrence of grade 3 adverse events associated with vaccination in 10% of participants or more.  Study early termination criteria:  - A grade 4 adverse reaction or serious adverse event that may be related to vaccination during the study period, and a decision to terminate the trial after discussion by an expert panel organized by the principal investigator.  - 15% or more of subjects with grade 3 or higher adverse reactions lasting >48 hours fail to reduce to grade 1 or 2, the principal investigator organizes an expert panel discussion to decide whether to terminate the trial.  - Principal investigator request for full termination of the trial and explain the reasons.  - The ethics committee requested full termination of the trial and explain the reasons.  - The administrative authority requested a full termination of the trial and explain the reasons. |
| Inclusion and  exclusion  criteria | Inclusion criteria.   - Healthy volunteers over 60 years of age 3-6 months after completing 2 doses (Group A and Group B) of basic immunization with inactivated SARS-CoV-2 vaccine, or 1-2 months after completing 1 dose (Group C and Group D) of inactivated SARS-CoV-2 vaccine. - Obtain informed consent from the subject and sign the informed consent form. - Subjects were able and willing to comply with the requirements of the clinical trial protocol and to complete the 6-month study follow-up. - Axillary temperature ≤37.0°C. - Subjects eligible for immunization with the products involved in this study were determined by medical history, physical examination, and clinical judgment.   Exclusion criteria.   - Those with a history or family history of convulsions, epilepsy, encephalopathy, and psychosis. - Allergy to any component of the study vaccine, history of more severe allergic reactions to vaccines, history of allergy - Women with a positive urine pregnancy test, who are pregnant, breastfeeding or planning to become pregnant within 6 months - People with acute febrile diseases and infectious diseases. - Have a serious chronic disease or a condition that is in a progressive stage that cannot be controlled smoothly, such as asthma, diabetes, thyroid disease, etc. - Congenital or acquired angioedema/neuroedema. - Have a history of urticaria 1 year before receiving the investigational vaccine. - Absence of spleen or functional absence of the spleen. - Thrombocytopenia or other coagulation disorders (which may cause contraindications for intramuscular injection). - Those who get dizzy from needles. - Have a history of immunosuppressive therapy, anti-allergy therapy, cytotoxic therapy, or inhaled corticosteroids (excluding corticosteroid spray therapy for allergic rhinitis, and acute corticosteroid therapy without dermatitis) over the past 6 months. - Received blood products within 4 months before injection of investigational vaccines. - Received another study drug within 1 month before injection of investigational vaccines - Received a live attenuated vaccine within 1 month before injection of investigational vaccines - Under anti-tuberculosis treatment. - In the judgment of the investigator, due to various medical, psychological, social, or other conditions that are contrary to the trial protocol or that affect the subject's ability to sign informed consent. |
| Principal Investigator | Name： Jingxin Li  Unit: Jiangsu Provincial Center for Disease Control and Prevention  Address: Room 330, Building A, No. 172 Jiangsu Road, Nanjing  Postal Code: 210009  Phone: 13813865838  |
| Laboratory  main detection  sponsor 1  (responsible for  blood sample  processing,  ELISA antibody  and neutralizing  antibody  detection) | Responsible person: Du Pan  Unit: Nanjing Vazyme Biotech Co., Ltd.  Address: Building C1-2, Hongfeng Technology Park, Kechuang Road, Nanjing Economic and Technological Development Zone  Postal Code: 210000  Tel: 13598857057  |
| Laboratory  main detection  sponsor 2  (responsible for  the against live  virus antibody  detection in P3  laboratory) | Responsible person: Guo Xiling  Unit: Jiangsu Provincial Center for Diseases Control and  Prevention  Address: No.172 Jiangsu Road, Nanjing City  Postal Code: 210009  Tel: 025-83759424  |
| Laboratory  main detection  sponsor 3 (cell immune  response, B  cells, T cells and  other major  immune cell  population  differentiation  and antibody  spectrum  detection unit) | Responsible person: Qi Hai  Unit: Tsinghua University  Address: A107, Tsinghua University School of Medicine, Haidian District, Beijing  Postal Code.100084  Phone.13911059637  |

Table2. Study visit schedule and visit content for sequential immunization subjects

| Number of visits | V1 | V2 | V3 | V4 |
| --- | --- | --- | --- | --- |
| Visiting time point | Day 1 | V0+14 days | V0+28 days | V0+6 months |
| Visiting time window | (±3 days) | (+3 days) | (+ 4days) | (±15 days) |
| Informed Consent | ● |  |  |  |
| Registration of demographic information | ● |  |  |  |
| Physical examination and interrogation screening | ● |  |  |  |
| Randomly entered group | ● |  |  |  |
| Blood collection | ●(20ml) | ●(20ml) | ●(20ml) | ●(20ml) |
| Inoculation and 30-minute observation | ● |  |  |  |
| Security Visits (AR/AE) | ● | ● | ● |  |
| Serious adverse event (SAE) reporting※ | ● | ● | ● | ● |
| Issuance of Diary Cards | ● |  |  |  |
| Recycle Diary Cards, Issue Contact Cards |  | ● |  |  |
| Recycling Contact Cards |  |  | ● |  |
| Record the Vaccination and Visit Record | ● | ● | ● | ● |
| Documentation of combined medications/combined vaccines | ● | ● | ● | ● |

**1、Background and Principle**

#### 1.1 Pathogen

2019 Novel coronaviruses belong to the beta genus of coronaviruses with envelopes, round or oval particles, often polymorphic, and 60-140 nm in diameter. Their genetic characteristics are distinctly different from SARS-CoV and MERS-CoV. Chinese scientists found 88% homology with the gene sequences of two bats (bat-SL-CoVZC45 and bat-SL-CoVZXC21) in Zhoushan, China. The novel coronavirus discovered in Wuhan is the seventh coronavirus that can infect humans and has not been found in humans before.

Coronaviruses belong to the genus Coronavirus in the family Coronaviridae in terms of phylogenetic classification. Viruses of the genus Coronavirus are positive-stranded single-stranded RNA viruses with an outer membrane and are a large group of viruses that are widely found in nature. Globally, 10%-30% of upper respiratory infections are caused by four types of coronaviruses, HCoV-229E, HCoV-OC43, HCoV-NL63, and HCoV-HKU1, which are the second leading cause of the common cold, after rhinovirus. The Middle East respiratory syndrome (MERS) and severe acute respiratory syndrome (SARS), which are known to be caused by coronaviruses, are serious infectious diseases.

The coronavirus genome sequentially encodes spike protein (S), an envelope protein (E), membrane protein (M), and nucleoprotein (N). The S protein is the most important surface protein of coronaviruses and is associated with the infectivity of the virus. The S protein has been used as the most important candidate antigen during the development of previous SARS and MERS vaccines.

#### 1.2 Disease and epidemiological background

The main manifestations of the disease are fever, dry cough, and weakness. A small number of patients have symptoms such as nasal congestion, runny nose, sore throat, myalgia, and diarrhea. Severe patients tend to develop respiratory distress and/or hypoxemia a week after the onset of the disease, which can rapidly progress to acute respiratory distress syndrome, septic shock, uncorrectable metabolic acidosis, bleeding, coagulation dysfunction, and multi-organ failure in severe cases. It is worth noting that heavy and critically ill patients may have low to moderate fever or even no significant fever during the disease. Some children and newborns may have atypical symptoms, such as diarrhea, vomiting, and other gastrointestinal symptoms, or they may only show weakness and shortness of breath. Patients with milder forms of the disease only show low fever and mild malaise, without pneumonia. From the current cases, most patients have a good prognosis, while a few patients are in critical condition. The prognosis is poorer in the elderly and those with chronic underlying diseases. The clinical course of pregnant women with novel coronavirus pneumonia is similar to that of patients of the same age. Childhood cases have relatively mild symptoms.

The current source of infection seen is mainly patients with novel coronavirus infection. Asymptomatic infected persons may also be the source of infection. Respiratory droplet and close contact transmission are the main routes of transmission. The potential for aerosol transmission exists in relatively closed environments with prolonged exposure to high aerosol concentrations. Novel coronaviruses can be isolated in feces and urine, and attention should be paid to aerosol or contact transmission due to environmental contamination by feces and urine. The human population is generally susceptible.

#### 1.3 Basis of the study

There are five main global 2019-nCoV vaccine development technology routes, namely inactivated vaccine, live attenuated vaccine, recombinant protein vaccine, viral vector vaccine, and nucleic acid vaccine. The inactivated vaccines of Beijing Sinovac Research & Development Co., Ltd. and the recombinant novel coronavirus vaccine (adenovirus vector) jointly developed by the Military Academy of Military Medical Institute and CanSino Biologics Inc. have been conditionally listed. The results of foreign phase III initial clinical trials showed that the short-term protection rate of the new inactivated coronavirus vaccine and the Ad5 vector novel coronavirus vaccine against symptomatic COVID-19 ranged from 50% to 70%, which was moderate, although it met the minimum 50% protection rate requirement proposed by the World Health Organization (WHO). There was no significant advantage over the 95% protection rates of Moderna and Pfizer/BioNTech mRNA vaccines.

To induce a sufficiently high level of the immune response, Russia was the first country in the world to adopt an immunization program of one dose of basal + one dose of booster immunization with a heterologous vector vaccine by using two different viral vectors of the 2019-nCoV vaccine, with a protection rate of 91.4% for the rAd26/rAd5 adenovirus vector vaccine. The UK has also announced a sequential vaccination study of adenoviral vector 2019-nCoV vaccine and mRNA vaccine, to optimize the existing immunization schedule of the respective 2019-nCoV vaccines and achieve better protection in a short period. In addition, with the global prevalence of SARS-CoV-2 variant strains, which poses a great challenge for the prevention of variant strains with a generation vaccine against the original strain, sequential immunization with a second-generation 2019-nCoV vaccine based on immunization with a generation 2019-nCoV vaccine offers the possibility to deal with SARS-CoV-2 variant strains.

Theoretically, there are significant differences in the types and characteristics of immune responses induced by the Ad5 vector 2019-nCoV vaccine and the inactivated 2019-nCoV vaccine, and sequential vaccination of the two vaccines may form a complementary advantage, improve the speed of immune response generation and the quality and immune persistence of the immune response, and optimize the existing immunization strategy of the 2019-nCoV vaccine.

This study is intended to evaluate the immunogenicity and safety of sequential immunization with inactivated vaccine and 2019-nCoV recombinant vaccine (Ad5 Vector) in a sequential immunization program. The clinical study protocol was developed following the requirements of the Vaccine Control Law, the Good Clinical Practice (GCP), the Technical Guidelines for Clinical Trials of Vaccines, and the Guidelines for Quality Management of Clinical Trials of Vaccines (Trial).

### 2 Purpose of the Study

Safety and immunogenicity of heterologous prime-boost immunization of Inactivated SARS-CoV-2 vaccine (Vero cells) and Recombinant COVID-19 vaccine (Ad5 Vector) in healthy adults aged 60 years and older in China.

### 3 Study Design

This study is a single-center, randomized, observer-blinded, parallel-controlled, sequential immune clinical trial.

#### 3.1 Study endpoint indicators

##### 3.1.1 Main endpoint indicators

- Incidence of adverse reactions within 28 days after sequential immunization in each group.
- Neutralizing antibodies to live SARS-CoV-2 (GMT) at 14day after sequential immunization in each group.

##### 3.1.2 Secondary endpoint indicators

1. Security endpoint

- Incidence of recruiting adverse events on day 0-14 after sequential immunization in each group.
- Incidence of non-recruiting adverse events on day 0-28 after sequential immunization in each group.
- Incidence of serious adverse events within 6 months after sequential immunization in each group.

2. Humoral immune study endpoints

- Geometric mean titers (GMT) of anti-SARS-CoV-2 S and N protein-specific antibody (ELISA) at Day14, Day28, and Month 6 after sequential immunization in each group.
- GMT of neutralizing antibodies to live SARS-CoV-2 at day 28 and months 6 after sequential immunization in each group.
- The Geometric Mean of the Fold Increase (GMFI) of antibody level of anti-SARS-CoV-2 S and N protein-specific antibody (ELISA) at Day14, Day28 and Month 6 after sequential immunization compared with Day0 before sequential immunization in each group
- The Geometric Mean of the Fold Increase (GMFI) of neutralizing antibodies to live SARS-CoV-2 at Day14, Day28, and Month 6 after sequential immunization compares with Day0 before sequential immunization in each group.
- The positive conversion rate of antibody level of anti-SARS-CoV-2 S and N protein-specific antibody (ELISA) at Day14, Day28, and Month 6 after sequential immunization compared with Day0 before sequential immunization in each group (≥ 4-fold increase).
- Positive conversion rates of neutralizing antibodies to live SARS-CoV-2 at Day14, Day28, and Month 6 after sequential immunization compare with Day0 before sequential immunization in each group (≥4-fold increase).

3. Cellular immunology study endpoints

- Levels of IFN-γ, TNF-α, IL-5, IL-4, and IL-13 were secreted by specific T cells at Day14 after sequential exemption in each group.

##### 3.1.3 Exploratory endpoint indicators

- Types of IgG antibodies against SARS-CoV-2 S protein at Day14, Day28, and Month 6 after sequential immunization in each group.
- Cross-neutralization of anti-SARS-CoV-2 neutralizing antibodies produced after sequential immunization of each group with other coronaviruses.
- Differentiation of major immune cell populations such as B cells and T cells and antibody profiles at Day14, Day28, and Month 6 after sequential immunization in each group.

#### 3.2 Estimation of sample size

（1）Hypothesis 1：GMT of Group A is not inferior to that in group B at Day28 after the boost vaccination.

（2）Hypothesis 2: GMT of Group A is superior to that in group B at Day28 after the boost vaccination.

*If hypothesis 1 holds, further statistical inferences can be made about hypothesis 2.

*The comparison of GMT after immunization in group C and group D was used as the initial exploration content, and sample size estimation without statistical inference was done. Considering the recruitment difficulty of the population of Group C and Group D, it was set to half of the sample size of Group A and Group B.

Three to six months after two doses of inactivated vaccine, the baseline GMT level before the booster is expected to be about 1:40 (log10X=1.6), and increase to 1:80 (log10X =1.9) after one dose of inactivated vaccine. After receiving one dose of the recombinant COVID-19 vaccine (Ad5 vector) as a booster, the GMT is estimated to reach 1:160 (log10X =2.2). The standard deviation is about 40 (log10X=0.6), the sample size is calculated:

Hypothesis 1, one-sided α = 0.025 significance level, 90% study power, GMT ratio of group A/group B Non-inferiority Margin is 0.67 (log10X =-0.174), the ratio of group A and group B is 1:1, and the sample size is 35 per group.

Hypothesis 2, one-sided α = 0.025 significance level, and the ratio of group A and group B are 1:1. To ensure a 90% study power, a sample size of 98 per group could show that the GMT level of group A after immunization is better than that of group B.

Therefore, two hypotheses were considered to be satisfied, with a sample size of approximately 100 individuals each in the sequential booster immunization group (Group A) and the conventional booster immunization group (Group B), and approximately 50 individuals in the sequential immunization group (Group C) and the conventional immunization group (Group D). The total sample size is approximately 300 individuals.

Table 1. sample size of each study group

| Code | Grouping | Sample size | Basic immunity | Vaccines | Sequential immunization |
| --- | --- | --- | --- | --- | --- |
| A | Sequential booster immunization group | 100 | Completion of two doses of inactivated vaccine | Adenovirus vector vaccine (1 dose) | Sequential immunization after the second dose of inactivated vaccine 3~6 months |
| B | Conventional booster immunization group | 100 |  | Inactivated vaccine (1 dose) |  |
| C | Sequential immunization group | 50 | Completion of one dose of inactivated vaccine | Adenovirus vector vaccine (1 dose) | Sequential immunization 1~2 months after the first dose of inactivated vaccine |
| D | Conventional immunization group | 50 |  | Inactivated vaccine (1 dose) |  |
|  |  | 300 |  |  |  |

Note: Subjects in Groups A and B must have completed two doses of inactivated vaccine. Subjects in Group C and Group D must have completed one dose of inactivated vaccine.

#### 3.3 Research Plan

A total of 4 visits, at day 0 (before vaccination), and at Day14, Day28, and Month 6 after the boost vaccination.

Table 2: Study visit schedule and visit content for sequential immunization subjects

| Number of visits | V1 | V2 | V3 | V4 |
| --- | --- | --- | --- | --- |
| Visiting time point | Day 1 | V0+14 days | V0+28 days | V0+6 months |
| Visiting time window | (±3 days) | (+3 days) | (+ 4days) | (±15 days) |
| Informed Consent | ● |  |  |  |
| Registration of demographic information | ● |  |  |  |
| Physical examination and interrogation screening | ● |  |  |  |
| Randomly entered group | ● |  |  |  |
| Blood collection | ●(20ml) | ●(20ml) | ●(20ml) | ●(20ml) |
| Inoculation and 30-minute observation | ● |  |  |  |
| Security Visits (AR/AE) | ● | ● | ● |  |
| Serious adverse event (SAE) reporting※ | ● | ● | ● | ● |
| Issuance of Diary Cards | ● |  |  |  |
| Recycle Diary Cards, Issue Contact Cards |  | ● |  |  |
| Recycling Contact Cards |  |  | ● |  |
| Record the Vaccination and Visit Record | ● | ● | ● | ● |
| Documentation of combined medications/combined vaccines | ● | ● | ● | ● |

#### 3.4 Randomization and blinding

##### 3. 4.1 Randomization method

The study was randomized using stratified block group randomization in a 2:2:1:1 ratio. Subject randomization tables were generated by an independent randomization professional via SAS 9.4 or above and imported into the Interactive Response Technology (IRT) system, accessible only to authorized personnel. Subjects, investigators, and the sponsor's study management team were blinded throughout the trial. Authorized, non-blinded study center personnel have access to subject grouping information through the IRT system and use the study vaccine for the corresponding group based on the grouping information.

##### 3..4.2 Blind state maintenance

Safety observers, laboratory testers, and statistical analysts were blinded in this study. Vaccine administration, formulation, and vaccination personnel were unblinded, and confidentiality agreements were signed to ensure that unblinded reports and documentation were shared only with unblinded personnel. Unblinded personnel are not involved in field work other than vaccine preparation and vaccination, and all other study personnel remains blinded. The unblinded vaccine preparer covers the side wall of the vaccine syringe with a study number label after the vaccine has been prepared for blinding the subject.

##### 3..4.3 Unveiling the blind procedure

In the event of serious complications and adverse events during the trial that affects the choice of management measures, the investigator may, in the opinion of the investigator, need to know the group of subjects and may break the blinding on an emergency basis. Emergency blinding was performed by the principal investigator through a randomized system.

Blinding will be performed during the initial analysis of safety and immunogenicity at 28 days after vaccination for the completed immunization program, but subjects and safety observers will remain blinded.

#### 3.5 Investigational vaccine

##### 3..5.1 Recombinant COVID-19 vaccine (Ad5 Vector)

Recombinant COVID-19 vaccine (Ad5 Vector) is a liquid formulation made by inoculating HEK293SF-3F6 cells with replication-deficient human type 5 adenovirus expressing novel coronavirus S protein. After amplification and purification, the liquid preparation made by adding appropriate excipients is used to prevent the disease caused by SARS-COV-2 infection.

Active ingredient: Recombinant replication-deficient human type 5 adenovirus expressing the novel coronavirus S protein (5×10^10^ VP).

Excipients: mannitol, sucrose, sodium chloride, magnesium chloride, polysorbate 80, hydroxyethyl piperazine ethanesulfonic acid, glycerin.

Package: vial

Specification: 0.5 ml/bottle (5×10^10^ VP)

Shelf life: tentatively 12 months

Storage: Store and transport at 2-8°C.

Route of inoculation: Intramuscular injection (IM) at the lower margin of the lateral deltoid muscle of the upper arm.

Immunization procedure: single-dose vaccination.

##### 3..5.2 Inactivated SARS-CoV-2 vaccine (Vero cells)

Manufacturer: Beijing Sinovac Research & Development Co., Ltd.

Specification: 0.5ml/bottle, for commercial packaging, stored according to instructions.

Shelf life: tentative12 months

Storage: Store and transport at 2-8°C.

Route of inoculation: Intramuscular injection (IM) at the lower margin of the lateral deltoid muscle of the upper arm.

Immunization procedure: single-dose vaccination.

#### 3.6 Criteria for pausing or early termination

The principal investigator will organize an expert panel meeting to decide whether to terminate the clinical trial early if one of the following conditions occurs.

1) Occurrence of one or more ≥grade 4 adverse reactions or serious adverse events that may be associated with vaccination.
2) Occurrence of grade 3 adverse events associated with vaccination in 10% of participants or more.

Study early termination criteria:

1) A grade 4 adverse reaction or serious adverse event that may be related to vaccination during the study period, and a decision to terminate the trial after discussion by an expert panel organized by the principal investigator.
2) 15% or more of subjects with grade 3 or higher adverse reactions lasting >48 hours failing to reduce to grade 1 or 2, the principal investigator organizes an expert panel discussion to decide whether to terminate the trial.
3) The principal investigator requested for full termination of the trial and explain the reasons.
4) The ethics committee requested full termination of the trial and explain the reasons.
5) The administrative authority requested a full termination of the trial and explain the reasons.

### 4 PARTICIPANTS

#### 4.1 Participants selection

Healthy adults aged 60 years and older, who completed 1 or 2 doses of inactivated COVID-19 vaccine basic immunization, are recruited as subjects after full informed consent.

#### 4.2 Selection criteria

1) Healthy volunteers over 60 years of age 3-6 months after completing 2 doses (Group A and Group B) of basic immunization with inactivated SARS-CoV-2 vaccine, or 1-2 months after completing 1 dose (Group C and Group D) of inactivated SARS-CoV-2 vaccine.
2) Obtain informed consent from the subject and sign the informed consent form.
3) Subjects were able and willing to comply with the requirements of the clinical trial protocol and to complete the 6-month study follow-up.
4) Axillary temperature ≤37.0°C.
5) Subjects eligible for immunization with the products involved in this study were determined by medical history, physical examination, and clinical judgment.

#### 4.3 Exclusion criteria

1) Those with a history or family history of convulsions, epilepsy, encephalopathy, and psychosis.
2) Allergy to any component of the study vaccine, history of more severe allergic reactions to vaccines, history of allergy
3) Women with a positive urine pregnancy test, who are pregnant, breastfeeding or planning to become pregnant within 6 months
4) People with acute febrile diseases and infectious diseases.
5) Have a serious chronic disease or a condition that is in a progressive stage that cannot be controlled smoothly, such as asthma, diabetes, thyroid disease, etc.
6) Congenital or acquired angioedema/neuroedema.
7) Have a history of urticaria 1 year before receiving the investigational vaccine.
8) Absence of spleen or functional absence of the spleen.
9) Thrombocytopenia or other coagulation disorders (which may cause contraindications for intramuscular injection).
10) Those who get dizzy from needles.
11) Have a history of immunosuppressive therapy, anti-allergy therapy, cytotoxic therapy, or inhaled corticosteroids (excluding corticosteroid spray therapy for allergic rhinitis, and acute corticosteroid therapy without dermatitis) over the past 6 months.
12) Received blood products within 4 months before injection of investigational vaccines.
13) Received another study drug within 1 month before injection of investigational vaccines
14) Received a live attenuated vaccine within 1 month before injection of investigational vaccines
15) Under anti-tuberculosis treatment.
16) In the judgment of the investigator, due to various medical, psychological, social, or other conditions that are contrary to the trial protocol or that affect the subject's ability to sign informed consent.

#### 4.4 Withdraw from the study

Participants have the right to withdraw from the study at any time during the study period, and the investigator should record the reason for withdrawal:

1) Loss of contact and early termination of the study.
2) Subjects asked to withdraw for no reason.
3) Subjects requesting withdrawal for reasons unrelated to the study, such as long-term absence, relocation, etc., should record the specific reasons for withdrawal.
4) Subjects requesting withdrawal for study-related reasons, such as intolerance of adverse effects, intolerance of biospecimen collection, etc., should have specific reasons for withdrawal recorded, and investigators should follow up subjects who withdraw due to AE/SAE until the event is resolved.
5) A subject may, at his or her discretion, completely terminate the study, including discontinuing all study actions such as vaccination, biospecimen collection, and safety observations, and the study data before the subject's withdrawal may be used for analysis. If the subject prohibits the investigator from continuing to use all study data about him/her, all study data before the subject's withdrawal will not be available for analysis.
6) Subjects may also partially terminate the study at their discretion, such as stopping only the vaccination, or stopping only the biological specimen collection, etc. The rest of the study as specified in the protocol should continue to be completed.

#### 4.5 Complete the study

##### 4.5.1 Complete the safety data collection

Subjects who completed immunization were observed for safety at 28 days post-immunization according to the protocol and for the occurrence of serious adverse events at 6 months post-immunization.

##### 4.5.2 Completion of Immunogenicity Study

Vaccination was administered according to the protocol, and follow-up blood collection at Day14, Day28, and Month 6 after vaccination was completed as specified in the protocol.

#### 4.6 Definition of violation and deviation from protocol events and measures taken

##### 4.6.1 Violation of protocol events

- Failure to provide informed consent to subjects and/or subjects not signing informed consent forms.
- Subjects did not meet the inclusion criteria/met the exclusion criteria and were enrolled in the study.
- Subjects received the wrong vaccine.
- Subjects who were administered a vaccine fail to meet the requirements.
- Any other reason as perceived by the investigator and confirmed by the principal investigator.

##### 4.6.2 Deviation from program events

- Beyond the visiting time window.
- Poor subject compliance and subjects not completing blood sample collection.
- Failure to report serious adverse events (SAEs) promptly.
- Subjects using prohibited drugs (intramuscular, oral, or intravenous administration of systemic corticosteroid therapy ≥ 2 mg/kg/day for ≥ 14 days, or other immunosuppressive drugs).
- Other causes as perceived by the investigator and confirmed by the principal investigator.

Investigators/Quality Controllers should report protocol violations or deviations to the Principal Investigator (or Project Coordinator) by fax/email as soon as possible after discovery. For protocol violations, they should also be reported to the Ethics Committee.

### 5 Methods and procedures

#### 5.1 Participants selection

Healthy adults aged 60 years and older, who completed 1 or 2 doses of inactivated COVID-19 vaccine basic immunization, are recruited as subjects after full informed consent.

#### 5.2 Informed Consent

In obtaining and documenting informed consent, the investigator shall comply with the relevant regulations, the GCP, and the ethical principles outlined in the Declaration of Helsinki. Before the start of the study, the investigator shall obtain written approval/consent from the ethics review committee for the informed consent and other documents provided to the subjects.

Before participation in this clinical study, the investigator should address the contents of the informed consent form to the subject and/or his or her witnesses and should give the subject and/or his or her witnesses sufficient time to consult the details of the study before signing the informed consent form. When giving informed consent information to multiple individuals, each subject and/or witness should be allowed to ask the investigator questions individually before signing the informed consent form.

The investigator shall maintain the signed informed consent form for each subject and shall provide the subject with a copy of the signed and dated informed consent form.

#### 5.3 Physical examination and screening

Subjects were required to undergo a physical examination, including temperature, and a urine pregnancy test for non-menopausal women before enrollment. The physician will conduct a history and screening for enrollment according to the "inclusion and exclusion criteria", and those who pass the screening will be enrolled and participate in randomization.

#### 5. 4 Vaccine distribution and inoculation

A unique study number is assigned according to the order in which the screened subjects reach the randomized grouping room. The non-blinded vaccine preparation staff assigns the corresponding study vaccine according to the subject's study number, and after completing the vaccine preparation, the study number sticky note is attached to the side wall of the syringe and given to the vaccination nurse, who completes the vaccination.

The inoculation site should be equipped with emergency drugs such as epinephrine hydrochloride and simple ventilators, heart monitors, and other first-aid equipment.

##### 5..4.1 Immunization routes and immunization procedures

The site of vaccination for the study was the lateral deltoid muscle of the upper arm, and the route of vaccination was intramuscular injection.

Sequential booster immunization procedure: Subjects received one booster dose of Inactivated SARS-CoV-2 vaccine (Vero cells) or Recombinant COVID-19 vaccine (Ad5 Vector), respectively, at 3-6 months after the completion of 2 doses of inactivated vaccine immunization.

Sequential immunization procedure: Subjects received one booster dose of Inactivated SARS-CoV-2 vaccine (Vero cells) or Recombinant COVID-19 vaccine (Ad5 Vector), respectively, at 1-2 months after the completion of 1 dose of inactivated vaccine immunization.

Vaccines should be shaken well before use and should be used immediately after opening. If there are cracks, unclear or invalid labels, or abnormalities in the appearance of vaccines, they should not be used.

##### 5.4.2 Management of vaccines

1) Vaccine storage: The temperature of the vaccine storage place needs to be controlled within the range of 2-8℃, and freezing is strictly prevented; the storage temperature of the vaccine should be recorded twice every working day, and each time should be the same as long as possible.

2)Vaccine transportation: The partner is responsible for transporting the vaccine for the study from the production site to the cold storage of the clinical study site unit, and the vaccine management personnel of the study site unit will jointly check and sign the acceptance by the responsible research institution (Jiangsu Provincial Center for Disease Control and Prevention (Jiangsu Provincial Institute of Public Health)), including the vaccine transport temperature record (conforming to the vaccine cold chain temperature), inspection report (qualified) and whether the vaccine is damaged.

#### 5.5 Safety follow-up and evaluation

##### 5. 5.1 Safety observation

All subjects should be observed for immediate reactions at the vaccination site for 30-minute safety observation after vaccination. The trained researchers should systematically observe each subject, record the local and systemic reactions within 30 minutes, and record the severity.

Within 14 days after vaccination, the investigator will conduct an active safety follow-up by systematically observing subjects for adverse events and instructing them to fill out diary cards based on their self-signs and symptoms. From day 15 to day 28 after vaccination, adverse events will be observed by passively collecting adverse events reported by subjects. From day 28 to month 6 after vaccination, the occurrence of SAEs reported by the subjects was passively collected.

##### 5. 5.2 Safety observation content and indicators

In this clinical study, according to the Guidelines for Grading Standards for Adverse Events in Clinical Trials of Vaccines for Prophylaxis (NMPA [ 2019] No.102 ), the safety observation contents and indicators were graded as follows: (Table 3 - Table 4)

**Table 3 Grading table for inoculation site (local) adverse events**

| **Symptoms/Signs** | **Grade 1** | **Grade 2** | **Grade 3** | **Grade 4** |
| --- | --- | --- | --- | --- |
| Pain | Does not affect or slightly affects physical activity | Affects limb movement | Impact on daily life | Loss of basic self-care, or hospitalization |
| Hard knots*, swelling (optional)** # | Diameter 2.5 ~ <5 cm or area 6.25 ~ <25 cm^2^ and does not affect or slightly affect daily life | Diameter 5 ~ <10 cm or area 25 ~ <100 cm^2^ or affects daily life | Diameter ≥ 10 cm or area ≥ 100 cm^2^ or ulceration or secondary infection or phlebitis or aseptic abscess or wound drainage or severe impact on daily life | Abscesses, exfoliative dermatitis, dermal or deep tissue necrosis |
| Rash*, erythema (optional)** # | Diameter2.5 ~ <5 cm or area6.25 ~ <25 cm^2^ and does not affect or slightly affect daily life | Diameter 5 ~ <10 cm or area 25 ~ <100 cm^2^ or affects daily life | Diameter ≥ 10 cm or area ≥ 100 cm^2^ or ulceration or secondary infection or phlebitis or aseptic abscess or wound drainage or severe impact on daily life | Abscesses, exfoliative dermatitis, dermal or deep tissue necrosis |
| Pruritus | Itching at the inoculation site, relieved on its own or within 48h after treatment | Itching at the inoculation site, not relieved within 48 h after treatment | Impact on daily life | NA |
| Cellulitis | NA | Need for non-injectable treatment (e.g., oral antibacterial, antifungal, and antiviral drug therapy) | Need for intravenous treatment (e.g., intravenous antibacterial, antifungal, and antiviral drug therapy) | sepsis, or tissue necrosis, etc. |

Note: In ^*^addition to direct measurement of the diameter to grade the evaluation, the progressive change of the measurement should be recorded.

^**^The maximum measurement diameter or area should be used.

#The evaluation and grading of sclerosis and swelling, rash, and redness should be based on the functional class and actual measurements, and a higher graded-index should be selected.

**Table 4 Grading table for non-vaccination site (systemic) adverse events**

| **Organ system signs/symptoms** | **Grade 1** | **Grade 2** | **Grade 3** | **Grade 4** |
| --- | --- | --- | --- | --- |
| **Fever [Axillary temperature (℃)** | | | | |
| 14＞Age | 37.3~ <38.0 | 38.0~ <38.5 | 38.5~ <39.5 | ≥39.5, lasting more than 3days |
| **Gastrointestinal System** | | | | |
| Diarrhea | Mild or transient, 3~4 times/day, abnormal stool pattern, or mild diarrhea lasting less1 than a week | Moderate or persistent, 5~ 7 times/day, abnormal stool pattern, or diarrhea > 1weeks | > 7times/day, abnormal stool properties, or hemorrhagic diarrhea, upright hypotension, electrolyte imbalance, requiring intravenous fluids > 2L | Hypotensive shock, requiring  Inpatient treatment |
| Difficulty swallowing | Mild discomfort when swallowing | Restricted diet | Very restricted in eating, talking; unable to eat solid food | Cannot eat liquid food; requires intravenous nutrition |
| anorexia | Decreased appetite, but not reduced food intake | Decreased appetite, reduced food intake, but no significant weight loss | Decreased appetite and significant weight loss | Need for interventions (e.g., gastric tube feeding, parenteral nutrition) |
| vomiting | 1 to 2 times/24 hours without interfering with activities | 3~5 times/24 hours or activity limitation | ＞6 times/24 hours or require intravenous rehydration | Need for hospitalization or other means of nutrition due to hypotensive shock |
| Disgusting | Transient (24<hour) or intermittent with normal food intake | Persistent nausea leading to reduced food intake (24~48 hours) | Persistent nausea resulting in little to no food intake (> 48 hours) or need for intravenous rehydration | Life-threatening  (e.g. hypotensive shock) |
| **Musculoskeletal and connective tissue** | | | | |
| Muscle pain  (Non-inoculation site) | Does not interfere with daily activities | Slightly affects daily activities | Severe muscle pain that severely affects daily activities | Emergency or hospitalization |
| Arthritis | Mild pain with inflammation, erythema, or joint swelling that does not interfere with the function | Moderate pain with inflammation, erythema, or joint swelling; impairs function but does not interfere with daily activities | Severe pain with inflammation, erythema, or joint swelling that interferes with daily activities | Permanent and/or disabling joint injury |
| Joint pain | Mild pain that does not interfere with the function | Moderate pain; requires pain relievers and/or pain that interferes with function but does not interfere with daily activities | Severe pain; requires painkillers and/or pain interferes with daily activities | Dysfunctional pain |
| **Nervous System** | | | | |
| Headaches | Does not interfere with daily activities and does not require treatment | Transient, minor interference with daily activities, may require treatment or intervention | Seriously interferes with daily activities and requires treatment or intervention | Persistent, requiring emergency care or hospitalization |
| Fainting | Near syncope without loss of consciousness (e.g., aura syncope) | Loss of consciousness, but no treatment required | Loss of consciousness, requiring treatment or hospitalization | NA |
| **Spiritual System** | | | | |
| Insomnia | Mild difficulty falling asleep, not affecting or slightly affecting daily life | Moderate difficulty sleeping, affecting daily life | Severe difficulty sleeping, seriously affecting daily life, requiring treatment or hospitalization | NA |
| **Skin and subcutaneous tissue** | | | | |
| Pruritus at non-inoculated sites (no skin damage) | Mild itching, not affecting or slightly affecting daily life | Itching interferes with daily life | Itching that makes it impossible to carry out daily life | NA |
| Skin mucosa abnormalities | Erythema/itching/color change | Diffuse rash/papular rash/dryness/flaking | Herpetic/exudative/desquamative/ulcerative | Exfoliative dermatitis involving mucous membranes, or erythema multiforme, or suspected Stevens-Johnsons syndrome |
| **Respiratory System** | | | | |
| Cough | Transient, no treatment required | Persistent cough, effective treatment | Paroxysmal cough that cannot be controlled by treatment | Emergency or hospitalization |
| Immune System | | | | |
| Acute allergic reaction* | Localized urticaria (blisters), no treatment needed | Localized urticaria, requiring treatment or mild angioedema, not requiring treatment | Widespread urticaria or angioedema requiring treatment or mild bronchospasm | Anaphylaxis or life-threatening bronchospasm or laryngeal edema |
| **Other** | | | | |
| Fatigue, lethargy | Does not interfere with daily activities | Interference with normal daily activities | Severe disruption of daily activities and inability to work | Emergency or hospitalization |
| Non-inoculation site pain#  (Specify the site when reporting) | Mild pain with no or minor interference with daily activities | Pain interferes with daily life | Pain that prevents you from carrying out your daily life | Disabling pain, loss of basic self-care |

Note: *Refers to type I hypersensitivity reactions.

**# refers to pain at non-inoculated sites other than muscle pain, joint pain, and headache**

**Other general principles of adverse event classification.**

For clinical abnormalities not covered in the grading scale above, adverse events were assessed for intensity grading according to the following criteria.

| **Grade 1** | **Grade 2** | **Grade 3** | **Grade 4** | **Grade 5** |
| --- | --- | --- | --- | --- |
| Mild: Short-term (<48 h) or mild discomfort that does not interfere with activity and does not require treatment | Moderate: Mild or moderate restriction of movement that may require consultation, no or only mild treatment | Severe: Significant restriction of movement, requiring medical consultation and treatment, possibly hospitalization | Critical: potentially life-threatening, severely restricted activity, requiring supervised treatment | Death |

##### 5. 5.3 Outcome of adverse events

The outcome of adverse events included:1) recovery; 2) not yet recovered; 3) recovery but with sequelae; 4) death; 5) lost to follow-up.

##### 5. 5.4 Relationship between Adverse Events and Test Vaccines

5 = positively related: the adverse event should be the result of a temporary occurrence following immunization; and/or consistent with a known pattern of response to the test drug; unlikely to be produced by factors such as the subject's clinical status, interfering treatment, and concomitant therapy; and occurring immediately after immunization, or a positive reaction at the injection site.

4 = likely to be relevant: the adverse event should be the result of a temporary occurrence after vaccination; and/or consistent with a known pattern of reactions to the test drug; and unlikely to have arisen from factors such as the subject's clinical status, interventional treatment, and concomitant therapy.

3=possibly relevant: the adverse event should be the result of a temporary occurrence after vaccination; and/or consistent with the known pattern of reactions to the test drug; but potentially arising from factors such as the subject's clinical status, interventional therapy, and concomitant treatment.

2=possibly unrelated: adverse events are most likely to be related to factors such as the subject's clinical status, interventional therapy, and concomitant treatment, and are not a known pattern of response to the test drug.

1=Unrelated: adverse events were not related to vaccination and were associated with the clinical status of the subject, interventional treatment, and concomitant treatment.

##### 5. 5.5 Serious adverse reaction/event management

Adverse events: are unintended medical events that occur in subjects in clinical trials and are not necessarily causally related to the vaccine/vaccination.

Adverse reactions: Unintended or damaging reactions that occur during vaccination at the prescribed dose and procedure, usually related to vaccination.

Serious Adverse Event: is a medically important event, whether or not related to a clinical trial of a vaccine, that: 1) results in death; 2) is life-threatening; 3) results in hospitalization or prolonged hospitalization; 4) results in permanent or significant disability/loss of function; 5) results in congenital anomalies or birth defects; 6) results in other medically important events that could occur without treatment as listed above.

Suspected and Unanticipated Serious Adverse Reactions (SUSAR): Suspected adverse reactions are defined as harmful reactions unrelated to the purpose of the drug at any dose in a subject that is considered by analysis to be at least probably related to the drug; unanticipated refers to adverse reactions of nature, magnitude, consequence, or frequency that differ from the expected risk described in previous protocols or other relevant information (e.g., documents such as investigator's manuals and instructions).

Subjects should report to the investigator as soon as possible any clinically significant disease/event that develops after vaccination. Subjects should receive appropriate treatment at the designated hospital following the relevant national regulations. The investigator should follow up on the adverse reaction/event until the symptoms disappear or the symptoms stabilize. If deemed necessary by the investigator, the necessary treatment and management will be provided unconditionally to relieve the subject of the suffering caused by the adverse reaction/event, and all medication and medical management will be recorded at each follow-up visit.

In the event of a serious adverse event/reaction, the investigator should take prompt action as necessary and report to the principal investigator within 24 hours.

##### 5. 5.6 Reporting procedures for serious adverse events

Any serious adverse event, whether or not related to the test vaccine, must be reported to the principal investigator by fax or e-mail by completing a Serious Adverse Event Report Form within 24 hours of notification: description of the adverse event, time, and type of episode, duration, intensity, a causal relationship to vaccination, outcome, management (symptomatic treatment), and other relevant clinical and Laboratory data.

When a serious adverse event is reported to the principal investigator, the principal investigator, together with the safety observer, should consider the duration, extent, intensity, regression, and the subject's wishes to decide whether the subject should continue to participate in the trial or terminate the trial early. The principal investigator is required to determine whether the SAE is a suspected and unanticipated serious adverse reaction (SUSAR), and if so, the principal investigator is required to promptly report it to the vaccine manufacturer, the General Administration Drug Review Center, and the State Health and Welfare Commission.

For suspected and unanticipated serious adverse reactions (SUSAR) that are fatal or life-threatening, the principal investigator should report to the Drug Review Center of the General Administration as soon as possible within 7 natural days after first being informed, and within the following 8 days to improve follow-up information.

Note: The day the principal investigator was first informed was 0day one.

For suspected and unanticipated serious adverse reactions that are not fatal or life-threatening, or other potentially serious safety risk information, the principal investigator should report to the General Administration Drug Review Center as soon as possible, but within 15 natural days, after first becoming aware. Investigators should document serious adverse events and evaluate and discuss them in the final report after the trial is completed or terminated.

##### 5. 5.7 Clinical Assessment

The investigator should report any clinically manifest adverse event after being informed of the subject's vaccination as soon as possible, promptly investigate and medically visit the adverse event, such as medical history, physical examination, and necessary laboratory tests, and perform appropriate medical management. For serious adverse reactions/events, follow-up should be continued until the serious adverse event is resolved and a detailed investigation and follow-up record are finally completed, including the following:

1) description of adverse events.

2) the start time and end time of the adverse event.

3) intensity grading.

4) correlation with vaccination.

5) laboratory test results.

6) processing measures.

##### 5. 5.8 Treatment of pregnancy events

Pregnancy is not allowed to participate in this trial vaccination. A urine pregnancy test will be performed on the subject before vaccination, and those with a positive urine pregnancy test will not be allowed to enter the group. If a pregnancy event occurs within 6 months of the visit, a Pregnancy Event Report Form will be completed.

##### 5..5.9 Combination of medications/vaccines

If a subject has a medical event during the study, the appropriate treatment and medical management are permitted, but it should be promptly documented what medications were used or what medical management was performed.

Subjects are not recommended to receive other vaccines during the study period, except for those requiring emergency vaccinations due to an emergency, including rabies vaccine, tetanus vaccine, or other vaccines requiring emergency vaccinations. Subjects will be required to keep detailed records of whatever vaccines they receive during the study period.

#### 5. 6 Sample collection and site management

##### 5. 6.1 Samples collection

For each visit of V1-V4, 20 ml of venous blood was collected by vacuum anticoagulated blood collection tubes, and PBMC and serum were separated to detect the level of vaccine-induced antibodies and the differentiation of immune cell populations and antibody profiles.

Blood specimens from this clinical trial will be used for immune response index testing as specified in the protocol, and approval from the ethics committee is required for use in other studies.

##### 5. 6.2 Preservation and transport of samples

Uniform operating standards are used for the storage and transportation process. The serum should be stored at -20°C and below and transported to the testing laboratory promptly. BPMC for immune cell population differentiation and antibody profiling assays are separated, transported, and stored by third-party laboratories according to standard operating procedures.

**6 Data administration**

#### 6.1 Data administration

In this study, EDC was used for the collection and management of study data, and the system retained the complete modification track to ensure the traceability of clinical trial data; the collection, entry, cleaning, consistency verification, and database locking of data were completed according to the relevant requirements of the Technical Guidelines for Clinical Trial Data Management. The data management process should conform to GCP specifications to ensure the authenticity, integrity, and accuracy of clinical trial data.

**6.1.1 Design and establishment of the database**

The creation of the project database (eCRF) is carried out by the database designer, and the database is created using the CDISC standard as much as possible.

After the database is established and tested and completed, each role authority personnel PI, Sub-I, CRC, PM, CRA, DM, etc. can be formally applied online only after training.

**6.1.2 Data entry**

Trained data entry staff to complete online data entry promptly after completing the visit.

Investigators are required to approve the data on the eCRF to confirm that the data recorded in the eCRF are authentic. Once data entry is complete, any changes to the data will be explained (comments) and automatically recorded in the system.

**6.1.3 Verification of data records**

The QC staff is required to verify the data records entered into the EDC on a regular or occasional basis to ensure that all data entered is consistent with the original documentation. If there are inconsistencies, the QC staff needs to send a challenge to the investigator in the corresponding place in the EDC system, and the investigator needs to verify the original information and update the entry until the EDC system is filled out completely. Before locking the database, the QC should double-check the subject's original data and the necessary investigator signatures, etc.

**6.1.4 Data verification**

The data manager manages the challenge of trial data based on the Data Verification Plan (DVP).

When data is entered into the EDC system, if there is illogical data, the system will automatically verify and issue queries (Query); these Queries need to be reviewed and answered by the researcher or authorized personnel when the updated data makes the logical verification not valid, the Query will be automatically closed; the automatically closed Query can be reviewed by the DM, and when the problem is not resolved, the DM can manually Add a query and continue communication with the research center until the issue is resolved.

In addition to automatic system verification, queries verified by SAS programming or manually by the data manager can be manually added to the EDC system on top of the Query when clarification/verification/confirmation by the investigator is required.

Before data locking, the data management needs to ensure that all Query is cleaned up and the investigator completes an electronic signature on top of the EDC system. To ensure the integrity and accuracy of patient data.

**6.1.5 Medical Coding**

Medical coders perform medical coding work. Medical coding of non-solicitation adverse events. Adverse events will be coded according to the MedDRA (version 21.1 or higher) dictionary.

Any inability to code due to improper, inaccurate, or ambiguous medical terminology provided during the coding process can be challenged by the DM to the researcher online in real-time.

Medical review of medical codes is required before database lock.

**6.1.6 Database locking**

The data lock list is completed and based on the procedures for database locking, the data manager, statistical analyst, and principal investigator sign a written approval of the database lock document, which is exported by the data manager in the specified format and handed over to the statistician for statistical analysis. If there is definite evidence of the need to unlock the data after locking, the researcher and relevant personnel are required to sign the unlocking document.

**6.1.7 External data management**

Immunogenicity data is managed as external data, and data management reviews and consistently checks external data, etc.

**6.2 Statistical analysis**

**6. 2.1 Selection of analysis data set**

Safety evaluation data set (SS):

All subjects who received vaccination after randomization should be evaluated for safety. Data from protocol violations should not be excluded.

Immunogenicity evaluation dataset:

Full analysis data set (FAS): The FAS is the ideal subject population determined according to the ITT (intentional analysis) principle, and all enrolled and vaccinated subjects are included in the FAS set, regardless of whether they have positive baseline antibody levels and regardless of whether they are in serious violation of the trial protocol.

Protocol compliant dataset (PPS):

a subset of the FAS in which subjects are more adherent to the protocol has no major violations of the trial protocol during the study period, and all meet the inclusion/exclusion criteria.

In this trial, the conforming protocol data set (PPS) was used as the primary analysis set. However, the FAS is also analyzed, and any inconsistencies between the FAS and PPS analysis results are discussed in the report.

**6. 2.2 Statistical methods of data**

Statistical analysis was performed by first checking the number of completed cases and case shedding; then the analysis of demographics and each relevant characteristic at baseline at the time of case enrollment in each group was performed to examine the comparability between groups; the evaluation of vaccine effects included the determination of evaluation indexes and the comparison of effects between groups; the safety evaluation included the statistics of clinical adverse reactions/events.

Criteria for exclusion of cases: those who did not meet the criteria for enrollment; those who failed to follow up with data and information after vaccination; those who had severe missing information and data after randomization; subjects who met the criteria for the withdrawal but did not withdraw; subjects who received the wrong vaccination.

The analysis of safety in this trial was a mainly descriptive analysis of the incidence of adverse reactions/events, and the χ^2^ test could be used for comparison between groups, and Fisher's exact probability method was used when necessary. After vaccination, the number of local adverse events and the number of people were counted separately (conventional calculation method), and the number of people was calculated according to the highest severity in both arms, and the number of cases was calculated according to the cumulative number of local adverse events that occurred at the vaccination site. The analysis of antibody levels of immunogenicity indicators is subject to logarithmic transformation and should be expressed as GMT, standard deviation, median, maximum and minimum values, and 95% confidence interval, and the inverse cumulative distribution should be plotted. Comparisons of categorical indicators between groups, such as antibody-positive conversion rates were calculated as frequencies and percentages using the χ^2^ test, and Fisher's exact probability method was used when necessary. Statistical analysis of repeated measures information was used for test data from different time points.

All statistical calculations were processed with SAS 9.4 statistical analysis system, and statistical analysis tests were performed with a two-sided test, giving the test statistic and its corresponding P-value, which was calculated directly when Fisher's exact probability method was used, with P ≤ 0.05 as the criterion for a statistically significant difference.

##### 6.2.3 Initial Analysis

Initial analysis of safety and immunogenicity was performed 28 days after vaccination for the completed immunization program.

##### 6.2.4 Analysis Software

The tests were all analyzed using SAS software version 9.4 or above.

**7 Monitoring of clinical trials**

#### 7.1 Quality assurance and quality control

The site quality control is strictly under the relevant requirements of the Good Clinical Practice (GCP) for drug trials.

Investigators in some positions are qualified as physicians or above and are trained in the clinical protocol and all trial-related procedures, including information about the test vaccine, procedures for obtaining informed consent, operating procedures for each position, and procedures for reporting adverse reactions/events, before participation in this study.

The investigator reviews each subject's data at each stage of the clinical trial to ensure that the content of the clinical trial meets the requirements of the protocol and that the data obtained are complete and credible. The quality controller performs quality control of the entire clinical trial process.

All work on-site was carried out in strict accordance with the clinical trial site operation manual. Each subject recorded his or her Diary Card and was followed up with retrospective surveys by the investigator and reviewed to guide the completion of the Diary Card.

The QCs perform a thorough verification of the original data, and after training, the data entry of eCRF is performed by a dedicated person, using the double-entry method, which is done independently by two people.

Calibration or standardization of the instruments used in this clinical trial.

#### 7.2 Modification of clinical protocols

After this protocol has been approved by the Ethics Committee, if there are significant modifications in the implementation process, the Ethics Committee must be declared for approval before implementation. The investigator shall not perform any protocol deviation or change without the consent of the sponsor and prior review by the Ethics Committee (EC) and written approval.

Any changes to the protocol, whether significant or non-significant, require that they be in writing. Substantial protocol revisions that would affect the safety of subjects, the scope of the study, or the scientific quality of this study require EC approval.

#### 7.3 Scheme deviation

The investigator is required to perform this clinical trial under the clinical trial protocol approved by the Ethics Committee and in compliance with GCP regulations. During the trial, the investigator must not deviate from the protocol except to eliminate imminent harm to the subject.

The research center should record all protocol deviations in the subject's original data, including but not limited to the time of protocol deviation, time of discovery, description of the event, and measures taken. In case of serious protocol deviations, the principal investigator should be notified and the ethics committee should be reported on time.

#### 7.4 Confidentiality

The investigator, the Ethics Committee (IEC), or a representative of a fully authorized regulatory body, has the right to access data related to the clinical trial, but its relevant content cannot be used in any other clinical trial or disclosed to any other person or entity.

The investigator must sign a confidentiality agreement to acknowledge that he or she knows and agrees to be responsible for the confidentiality of the information in this study.

The investigator and other study personnel shall maintain the confidentiality of all information provided by the sponsor, and all data/information generated at the study center (except medical records of subjects). This information and data may not be used for any purpose other than the study. This restriction does not apply to (1) research information that is not released as a result of a breach by the investigator and investigator; (2) research information that is released to the IRB/IEC for research evaluation purposes only; (3) research information that is released to provide appropriate medical assistance to subjects.

**7.5 Quality control of documentation**

**7. 5.1 Original information**

1) original record book.

2) informed consent form.

3) specimen collection records.

4) vaccination records.

5) adverse event observation results and records.

6) cold chain records.

7) Vaccine handover, use, distribution, and recall records.

**7.5.2 Preservation of information**

Data from clinical trials shall be preserved under Appendix 2 of the GCP, and the investigator shall retain the study data up to five years after the termination of the clinical trial.

**7.6 Quality control of biological sample**

**7. 6.1 Quality control of biological sample collection**

Blood collection using 10ml vacuum anticoagulated blood collection tubes. The sampler should verify the basic information and process in the original record book, and the blood should be locally disinfected before blood collection. After specimen collection, the subject ID and specimen number must be indicated in the relevant documents and blood collection tubes and checked correctly on the spot. After blood collection and numbering, the sampler should sign in the appropriate place in the original record book. Special circumstances of blood collection should be accurately recorded.

There is a person responsible for the quality check of the blood collection process, specimen quality, and document filling. In case of a wrong number, heavy number, or specimen failure, the person in charge of the site should be contacted immediately for a timely remedy.

The collected specimens should be properly stored, handed over to the laboratory blood sorting personnel promptly and handover records should be made, and medical waste should be sorted and placed as required and handed over to the relevant person in charge for disposal on time.

**7.6.****2 Quality control of biological samples during transportation**

Field specimens need to be transported to the specimen testing laboratory, and all specimens need to be recorded under the field standard operating regulations.

The site logistics manager needs to compile the specimen shipping sheet before sending the specimens: the content should include the number of specimens, specimen box number, specimen number, etc. A paper version of the specimen shipping slip is shipped with the specimens.

Upon receipt of specimens, the laboratory specimen recipient should count the number and condition of the specimens, check whether the specimens and the shipping order are consistent, check whether the specimen number is unique, and sign the specimen shipping order.

There should be temperature monitoring records during the transportation of serum specimens.

**7.6.3 Quality control of biological sample preservation**

The temperature of all project-related refrigerators should be monitored1 morning and evening, and if the temperature is abnormal, the person in charge of the refrigerator needs to fill in the temperature monitoring form with the reason for the abnormality and the treatment measures. Including cold chain interruption alarm and other processing.

**8 Risk Management Plan**

**8.1 Safety specifications**

Includes important identified risks, important potential risks, and important missing information. A comprehensive consideration is made based on the ADRs collected and summarized in previous clinical studies, the pharmacokinetic-pharmacokinetic characteristics of the product, the risk of medical treatment/intervention required, class reactions, epidemiology of the indication and target population, safety risks observed in non-clinical trials (including toxicology, drug interactions, etc.), and populations not studied in clinical trials.

**8.2 Pharmacovigilance programs**

Refer to the data, literature, or reports of similar vaccine safety-related studies at home and abroad, closely monitor and report SUSAR and potential safety risks occurring in the study vaccine; if significant safety risk warnings or reports are available, a risk control plan should be formulated and necessary measures are taken to protect the safety of the subjects. Safety monitoring information in clinical trials should be regularly summarized and analyzed following relevant requirements. In the analysis of the monitoring data, focus on the safety risk signal of the drug. Based on the analysis of the monitoring data, further, identify the differences between the monitoring data and the safety information in the drug instructions, analyze the occurrence of new and serious adverse reactions, discuss the need for risk management measures and provide advice on benefit-risk assessment.

**8.3 Risk minimization measures**

The main measures include: updating and revising the entry criteria and informed consent form of the study protocol promptly based on the information collected through long-term observation and follow-up; additional risk minimization measures including risk grading of identified risks and issuing recommendations for treatment measures, strengthening communication with subjects and giving appropriate training to participating study personnel to convey recommendations for treatment related to risk grading, etc. In the trial protocol, uniform safety evaluation standards and methods are developed regarding the corresponding guidelines issued by the National Bureau, and the safety of the vaccine is proactively monitored and followed up.

**9 Schedule**

Total study duration: This clinical trial is planned to last approximately 6 months.

**10 Ethics Approval**

This clinical trial protocol shall be approved by the Ethics Review Committee. The principal investigator submits the clinical protocol and all necessary additional documents to the ethics committee, and after the ethics committee approves and agrees, the Ethics Review Approval is issued to the investigator.

The investigator will also be required to provide a sample informed consent form to the ethics committee and have it approved by the ethics committee.

Subjects were given sufficient time to consider whether to participate in this trial before signing the informed consent form. Subjects were allowed to ask questions about the details of the trial and receive detailed answers. During the trial, subjects have the right to decide whether to withdraw from the trial.

**10.1 Ethical review and approval**

The principal investigator submits the clinical trial protocol and all necessary additional documents to the ethics committee for initial review.

- The clinical study protocol (indicate version number/version date)
- Informed consent form (specify version number/version date)
- Materials for subject recruitment (indicate version number/version date)
- eCRF sample (with version number/version date)
- Diary card (with version number/version date)
- Vaccination visit record (indicate version number/version date)
- Principal Investigator's Professional Biography
- Research vaccine inspection reports or batch release documents

**10.2 Supervision of the following implementation processes**

**10.2.1 Informed Consent**

Whether the method of subject enrollment and the provision of relevant information materials to subjects are complete and easy to understand; and whether the method of obtaining informed consent is appropriate. Throughout the trial, the ethics committee monitors whether there are ethical concerns about harm to subjects and whether subjects are treated or compensated for any harm caused by the trial, and evaluates the degree of risk to subjects.

**10.2.2 Confidentiality**

Ensure subject personal confidentiality under the conditions of test conduct and biological specimen collection as well as reporting, publication, etc. Only ID numbers, random numbers, and specimen numbers are recorded for biological samples.

**10.2.3 Potential risks and minimization of risks**

##### 10.2.3.1 Benefits and Risks

Subjects will likely receive better immune protection against the SARS-COV-2 virus with a booster dose of the vaccine or completion of routine or sequential immunization with inactivated SARS-COV-2 vaccine. Since both Inactivated SARS-CoV-2 vaccine (Vero cells) and Recombinant COVID-19 vaccine (Ad5 Vector) have demonstrated a better safety profile in mass vaccination, an additional dose of vaccine is not expected to pose a significantly increased safety risk. Subjects participating in this clinical trial will not be required to pay for a vaccination with the investigational vaccine. Subjects will receive reasonable compensation for transportation, lost wages, blood collection, and nutrition for participation in this trial.

Vaccine injections may cause some adverse reactions. Common adverse reactions to vaccination include fever, injection site tenderness, redness, and swelling. Usually, the adverse reactions will resolve within 3-5 days after their appearance. Rapid and severe allergic reactions after vaccination are very rare but can be life-threatening. Therefore, during your stay at the clinic after vaccination, your health condition will be observed and evaluated by specialized medical staff. If an allergic reaction occurs, symptomatic treatment will be given at the first opportunity.

In addition, petechiae and mild pain at the site of blood collection may occur during blood collection. Although fainting during blood collection and infection at the collection site is very rare, they can occur.

##### 10.3.2.2 Vaccination

Procure regular and qualified vaccination consumables and conduct aseptic vaccination in strict accordance with standard methods to avoid adverse events caused by improper vaccination or vaccination errors.

If a subject has a Grade 3 or higher adverse reaction during the safety observation period, or if an SAE related or possibly related to the test vaccination occurs, he or she should be able to receive prompt medical treatment, and if necessary, the green channel for medical treatment should be immediately activated for emergency treatment.

##### 10.3.2.3 Blood specimen collection

After review of the principal investigator's qualifications, experienced nursing staff was hired and trained to perform venous blood specimen collection according to prescribed procedures to minimize the amount of pain or danger (including pain and, rarely, venipuncture site infection) experienced by the subject.

**11 Data Disclosure and Publication**

After the completion of this clinical trial, if the results of the trial are to be disclosed and/or published, the positive results will be disclosed and/or published together with the negative results.
